# Effectiveness of *Wolbachia*-infected mosquito deployments in reducing the incidence of dengue and other *Aedes*-borne diseases in Niterói, Brazil: a quasi-experimental study

**DOI:** 10.1101/2021.01.31.21250726

**Authors:** Sofia B. Pinto, Thais I. S. Riback, Gabriel Sylvestre, Guilherme Costa, Julia Peixoto, Fernando B. S. Dias, Stephanie K. Tanamas, Cameron P. Simmons, Suzanne M. Dufault, Peter A. Ryan, Scott L. O’Neill, Frederico C. Muzzi, Simon Kutcher, Jacqui Montgomery, Benjamin R. Green, Ruth Smithyman, Ana Eppinghaus, Valeria Saraceni, Betina Durovni, Katherine L. Anders, Luciano A. Moreira

## Abstract

**Background:** The introduction of the bacterium *Wolbachia* (*w*Mel strain) into *Aedes aegypti* mosquitoes reduces their capacity to transmit dengue and other arboviruses. Evidence of a reduction in dengue case incidence following field releases of *w*Mel-infected *Ae. aegypti* has been reported previously from a cluster randomised controlled trial in Indonesia, and quasi-experimental studies in Indonesia and northern Australia.

**Methods:** Following pilot releases in 2015 – 2016 and a period of intensive community engagement, deployments of adult *w*Mel-infected *Ae. aegypti* mosquitoes were conducted in Niterói, Brazil during 2017 – 2019. Deployments were phased across four release zones, with a total area of 83 km^2^ and a residential population of approximately 373,000. A quasi-experimental design was used to evaluate the effectiveness of *w*Mel deployments in reducing dengue, chikungunya and Zika incidence. An untreated control zone was pre-defined, which was comparable to the intervention area in historical dengue trends. The *w*Mel intervention effect was estimated by controlled interrupted time series analysis of monthly dengue, chikungunya and Zika case notifications to the public health surveillance system before, during and after releases, from release zones and the control zone.

**Results:** Three years after commencement of releases, *w*Mel introgression into local *Ae. aegypti* populations was heterogeneous throughout Niterói, reaching a high prevalence (>80%) in the earliest release zone, and more moderate levels (prevalence 40 -70%) elsewhere. Despite this spatial heterogeneity in entomological outcomes, the *w*Mel intervention was associated with a 69% reduction in dengue incidence (95% confidence interval 54%, 79%), a 56% reduction in chikungunya incidence (95%CI 16%, 77%) and a 37% reduction in Zika incidence (95%CI 1%, 60%), in the aggregate release area compared with the pre-defined control area. This significant intervention effect on dengue was replicated across all four release zones, and in three of four zones for chikungunya, though not in individual release zones for Zika.

**Conclusions:** We demonstrate that *w*Mel *Wolbachia* can be successfully introgressed into *Ae. aegypti* populations in a large and complex urban setting, and that a significant public health benefit from reduced incidence of *Aedes*-borne disease accrues even where the prevalence of *w*Mel in local mosquito populations is moderate and spatially heterogeneous. These findings are consistent with the results of randomised and non-randomised field trials in Indonesia and northern Australia, and are supportive of the *Wolbachia* biocontrol method as a multivalent intervention against dengue, chikungunya and Zika.

## Introduction

Dengue is a mosquito-borne disease transmitted primarily by the *Aedes aegypti* mosquito, which has increased globally in both case burden and geographic footprint over the past 50 years. Approximately 40% of the world’s population are at risk of dengue transmission, with an estimated 400 million infections per year resulting in 50 – 100 million clinical cases and 3.6 million hospitalisations.^1,2^ The economic cost to health systems and communities has been estimated at $8.9 billion per annum.^3^ In Brazil, more than 1.5 million dengue cases and 782 deaths were reported nationally in 2019, with in excess of 1300 cases per 100,000 population in the worst affected Central-West region. In the same year 132,000 cases of chikungunya - also transmitted by *Ae. aegypti* mosquitoes - were reported, including 92 deaths.

Current strategies for dengue control are limited to efforts to suppress immature and adult mosquito numbers, through spraying of insecticides and community campaigns to reduce breeding sites. Even where considerable resources are invested in these activities, sustained suppression of mosquito densities has been elusive, and seasonal outbreaks continue to occur.^4,5^ There is a well-recognised need for new, affordable and effective tools for control of dengue and other *Aedes*-borne arboviruses, including chikungunya and Zika.^4,6^

Stable introduction of the common insect bacterium *Wolbachia* (*w*Mel strain) into *Ae. aegypti* has been shown in the laboratory to result in *Ae. aegypti* having reduced transmission potential for dengue and other *Aedes*-borne arboviruses including chikungunya, Zika, Yellow Fever and Mayaro virus.^7-14^ Female *Ae. aegypti* mosquitoes infected with *w*Mel transmit the bacterium with high fidelity to their offspring via infected eggs and *w*Mel manipulates mosquito reproductive outcomes via a process called cytoplasmic incompatibility, which favours introgression of *w*Mel into a wild-type population.^13^ Accumulating evidence from field sites in Australia and Indonesia has demonstrated large reductions in dengue incidence in areas where short-term releases of *w*Mel-infected mosquitoes have resulted in introgression and sustained high prevalence of *w*Mel in local *Ae. aegypti* populations.^15-17^ A recently completed cluster randomised trial of *w*Mel *Wolbachia* deployments in Yogyakarta, Indonesia, conclusively demonstrated the efficacy of the method, with a 77% reduction in dengue incidence in *Wolbachia-*treated neighbourhoods compared to untreated areas.^18^ The Yogyakarta CRT included chikungunya and Zika as secondary endpoints, but insufficient cases were detected to permit an evaluation of efficacy against these arboviruses. Acquiring field evidence for the effectiveness of *Wolbachia* in reducing transmission of these arboviruses is a priority, as is the accumulation of real-world evidence for public health impact from large-scale implementations of *w*Mel-infected *Ae. aegypti* in the complex urban environments common throughout dengue-endemic areas.

Pilot releases of *Wolbachia*-infected mosquitoes started in 2014 in Rio de Janeiro and in 2015 in Niterói, Brazil, and achieved successful establishment of *Wolbachia* throughout the two small pilot site communities, each with a population of 2500-2800 people.^19,20^ In 2017 Niterói became the first site in Brazil to move to scaled deployments across a large urban area. The intervention involved a phased approach including engagement with and acceptance by the community, communication strategies to ensure the communities were informed and supportive, releases of *Wolbachia*-infected *Ae. aegypti* mosquitoes, and monitoring of the levels of *Wolbachia* in *Ae. aegypti* in the field.

We report here the entomological and epidemiological outcomes of a large-scale non-randomised deployment of *Wolbachia*-infected *Ae. aegypti* mosquitoes in the Brazilian city of Niterói, for the control of dengue and other *Aedes*-borne diseases. The impact of *Wolbachia* deployment on dengue, chikungunya and Zika incidence was evaluated via a quasi-experimental study, using controlled interrupted time series analysis of routine notifiable disease surveillance data, in accordance with a pre-defined protocol.^21^

## Methods

### Study setting

Niterói, a municipality of the state of Rio de Janeiro is situated in the Guanabara Bay across from Rio de Janeiro city (<u>22°52′58″S 43°06′14″W</u>). According to the last national census in 2010 it had a population of 484,918 living in an area of 135 km^2^. The city is divided into 7 health districts for administrative planning. For the evaluation of the impact of *Wolbachia* mosquito deployments, Niteroi was divided into four release zones and 1 control zone, which are aligned with neighbourhood administrative boundaries (Figure 1). Table 1 shows the baseline characteristics and release summary of each zone.

**Table 1:**
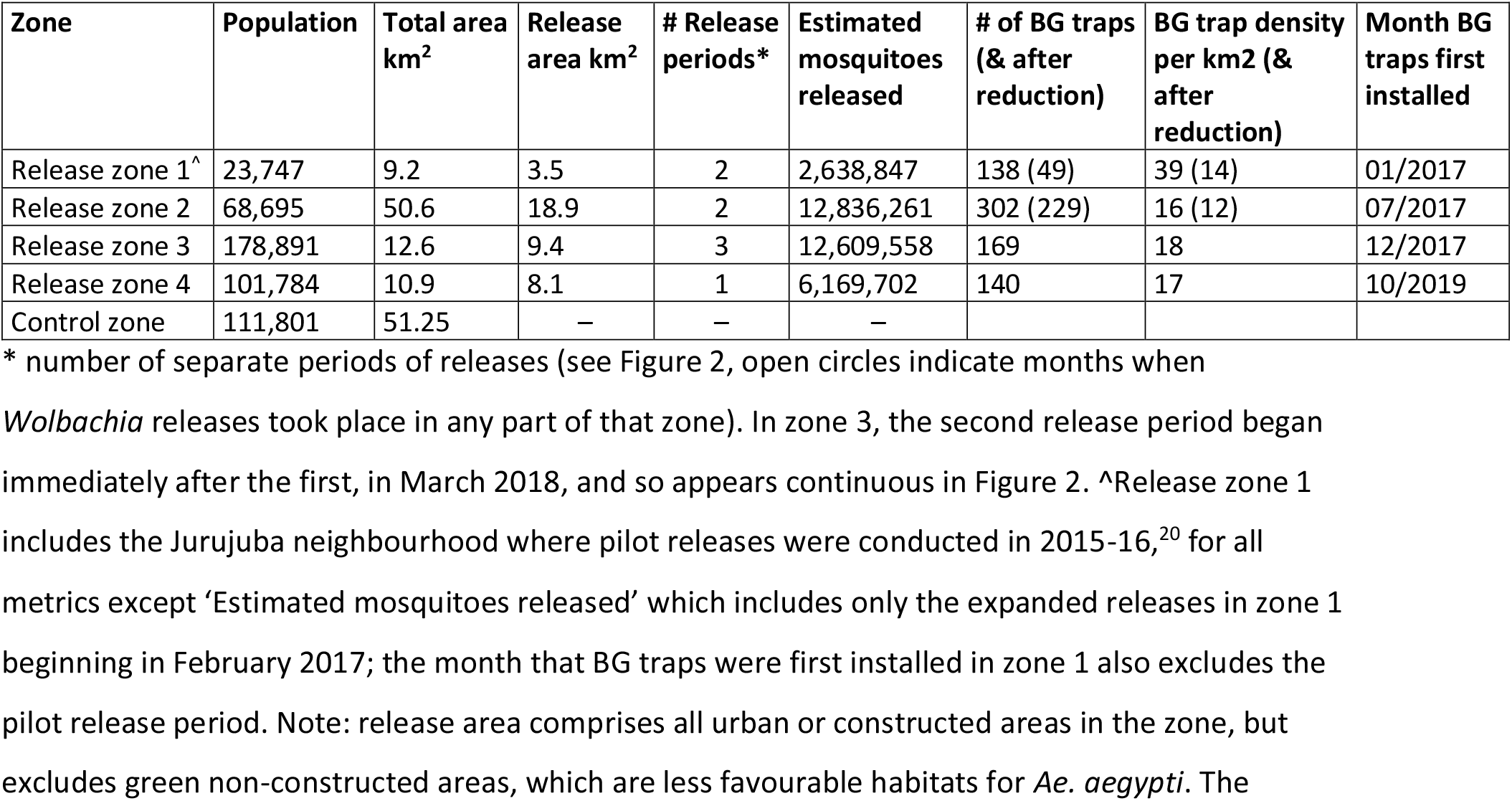
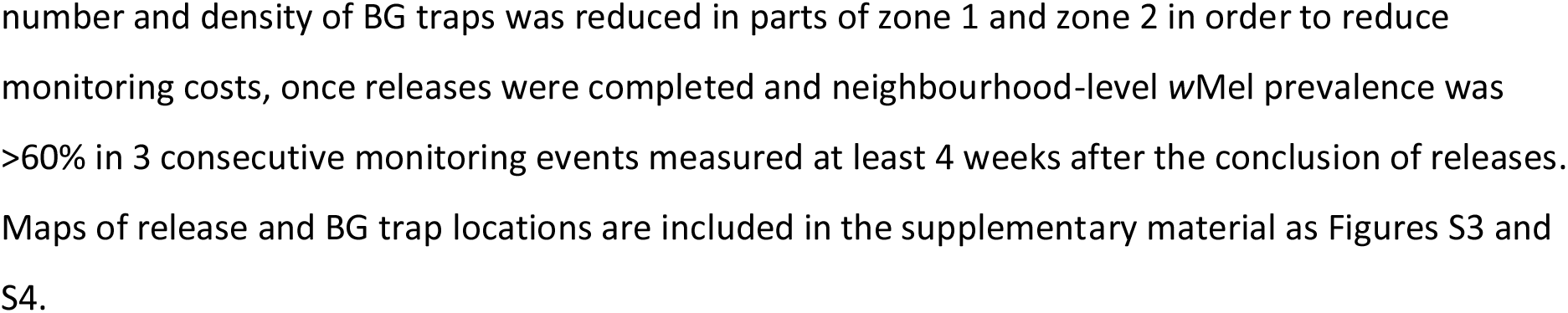
Baseline characteristics and summary of *w*Mel releases and monitoring by zone

| Zone | Population | Total area km<sup>2</sup> | Release area km<sup>2</sup> | # Release periods* | Estimated mosquitoes released | # of BG traps (& after reduction) | BG trap density per km<sup>2</sup> (& after reduction) | Month BG traps first installed |
| --- | --- | --- | --- | --- | --- | --- | --- | --- |
| Release zone 1<sup>^</sup> | 23,747 | 9.2 | 3.5 | 2 | 2,638,847 | 138 (49) | 39 (14) | 01/2017 |
| Release zone 2 | 68,695 | 50.6 | 18.9 | 2 | 12,836,261 | 302 (229) | 16 (12) | 07/2017 |
| Release zone 3 | 178,891 | 12.6 | 9.4 | 3 | 12,609,558 | 169 | 18 | 12/2017 |
| Release zone 4 | 101,784 | 10.9 | 8.1 | 1 | 6,169,702 | 140 | 17 | 10/2019 |
| Control zone | 111,801 | 51.25 | — | — | — |  |  |  |

\* number of separate periods of releases (see Figure 2, open circles indicate months when *Wolbachia* releases took place in any part of that zone). In zone 3, the second release period began immediately after the first, in March 2018, and so appears continuous in Figure 2. <sup>^</sup>Release zone 1 includes the Jurujuba neighbourhood where pilot releases were conducted in 2015-16,<sup>20</sup> for all metrics except ‘Estimated mosquitoes released’ which includes only the expanded releases in zone 1 beginning in February 2017; the month that BG traps were first installed in zone 1 also excludes the pilot release period. Note: release area comprises all urban or constructed areas in the zone, but excludes green non-constructed areas, which are less favourable habitats for *Ae. aegypti*. The

**Figure 1:**
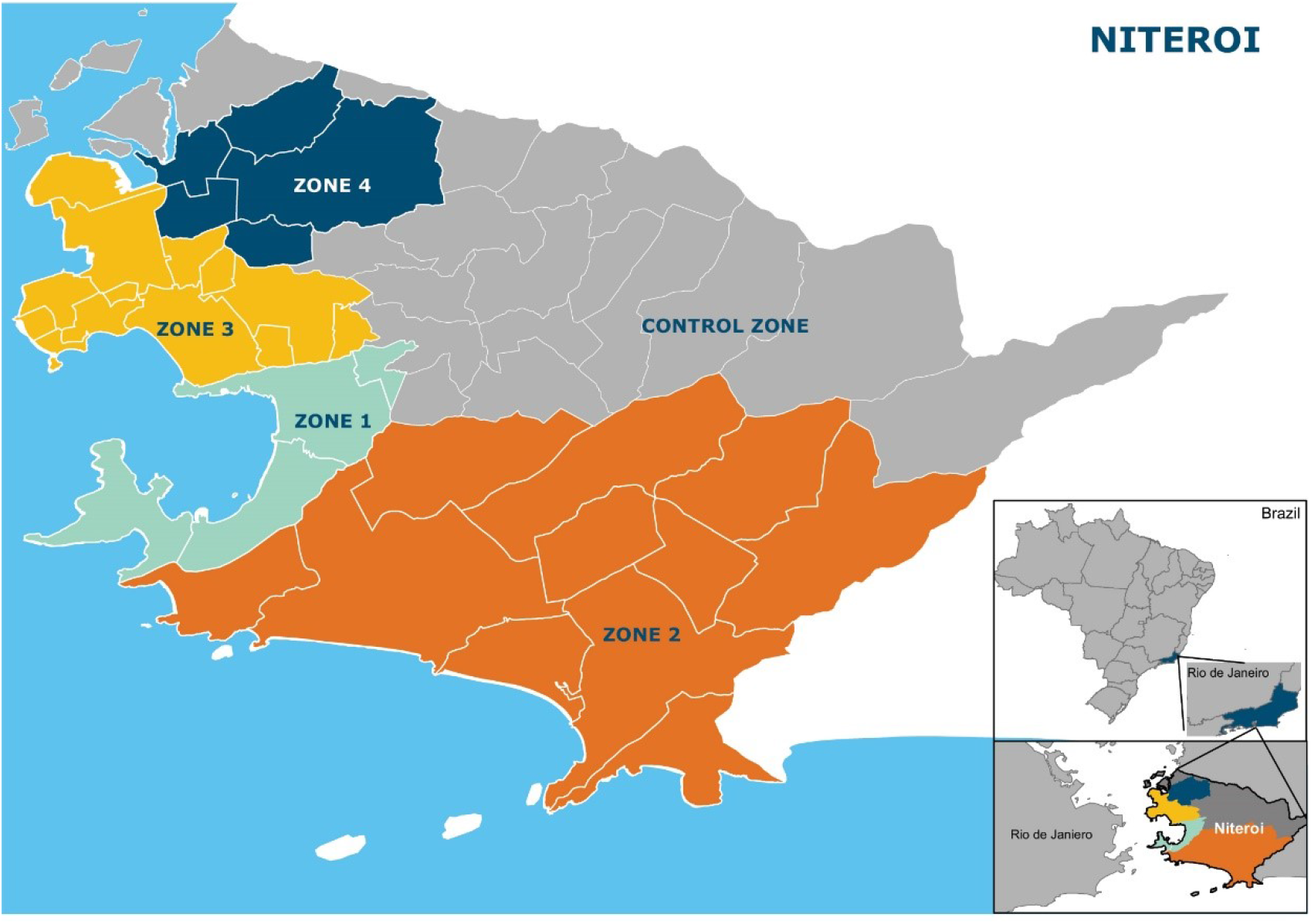
Study site map showing the municipality of Niterói, comprising four zones in which releases of *w*Mel-infected *Aedes aegypti* have been undertaken and one pre-defined parallel untreated control zone. Neighbourhood boundaries are shown in white. The inset shows the location of Niterói within the state of Rio de Janeiro, Brazil.

### Ethics and approvals

Approval to release *Wolbachia-*carrying *Ae. aegypti* mosquitoes into urban areas was obtained from three Brazilian governmental bodies: the National Agency of Sanitary Surveillance (ANVISA); the Ministry of Agriculture, Livestock and Supply (MAPA); and the Brazilian Institute of Environment and Renewable Natural Resources (IBAMA), which issued a Temporary Special Registry (Registro Especial Temporário (RET), nr. 0551716178/2017). Ethical approval was also obtained from the National Commission for Research Ethics (CONEP - CAAE 59175616.2.0000.0008).

### Community engagement

WMP Brazil’s Communication and Engagement (C&E) strategy was developed prior to mosquito releases, following a thorough analysis of geographical, social, political, economic and cultural factors in the proposed release areas as previously described.^22^

In Niterói the C&E plan was focused on three key areas: public schools, primary health care units and social leadership, due to their reach and influence within the release area, including into vulnerable communities. Community Reference Groups (CRGs) were also created, to serve as advisory committees populated by representatives of the planned release areas, to inform the activities of WMP Brazil. This group was also responsible for providing feedback on all communication materials and C&E strategies that were proposed throughout the WMP’s activities in their areas.

Prior to the release of *w*Mel-infected mosquitoes in each area, a survey of awareness and acceptance of the method was conducted by an independent company. In order to reach a wide range of people living and working in the release areas, time-location sampling was used to survey passers-by in busy public locations in each neighbourhood. Respondents (n= 3485 in total) were 18 years and over, and lived or worked in the neighbourhood where the survey was conducted. The questionnaire was developed with the CRG, and included questions on awareness (“Have you heard about the *Wolbachia* method?”), understanding after explanation of the method (“Do you understand that this method replaces the population of *Aedes aegypti* mosquitoes with *Aedes aegypti* mosquitoes carrying *Wolbachia*, which have a reduced capacity to transmit dengue, Zika and chikungunya?”) and acceptance of the proposed *w*Mel releases (“Do you agree with Fiocruz releasing these mosquitoes with *Wolbachia* here in your neighbourhood?”).

### Mosquito production

The Rio *w*Mel-infected *Ae. aegypti* line described in Garcia et al 2019^23^ was used for releases. The *w*Mel-infected lines were maintained in controlled laboratory conditions, in 900 cm^2^ mesh-sided rearing cages. Each cage contained 2500-2750 adults, and was fed using donated non-transfusional usable human blood (agreement FIOCRUZ/ Hemominas OF.GPO/CCO-Nr224/16), once per week for two to three gonotrophic cycles. As a quality assurance procedure each blood bag was tested for dengue, Zika, chikungunya, Mayaro and yellow fever viruses, as described previously^9,11,24^. Two separate colonies were maintained, a broodstock (kept in Belo Horizonte) and a release-production colony (kept in Rio de Janeiro). Male *Ae. aegypti* adults (from F0–F1 field collected material) were introduced into the broodstock cages at a rate of 10-20% every 5 generations. This outcrossing frequency was sufficient to maintain kdr resistant genotypes within the broodstock colony throughout its maintenance (see supplementary methods and Figure S1). Material from the broodstock colony was then transferred to the release-production colony where it was amplified through 2 amplifications without the addition of field collected males. A minimum sample of 168 mosquitoes from the release-production colony was screened for *w*Mel infection on a weekly basis, using quantitative polymerase chain reaction (qPCR) as described below. *w*Mel prevalence was 100% in all but three weekly screening events, and was never below 97%. Quantitative analysis of *w*Mel in these samples detected a fairly constant wsp:rps17 copy number between 4 to 6 (Figure S2).

From April 2017 until April 2018 immature stages for adult releases were reared at a density of approximately 1.0 larvae/ml and fed a diet of ground Tetramin Tropical Flakes (Tetra Holding [US] Inc. Germany, Product number 77101). From May 2018, immature stages for adult releases were reared at a density of approximately 2.75 larvae/ml and fed a diet of fish food: liver powder: yeast extract (4:3:1). We found no detrimental effects on outcomes, including development time, size, egg output or *w*Mel density, with increases in larval density up to 2.75/ml. In both rearing regimes, when approximately 10-30% of larvae had pupated, the larvae/pupae were sieved and between 180-220 larvae/pupae were placed in a release device. The release device was a cylindrical PVC crystal tube approximately 28 mm in diameter and 250 mm in length, covered with a fixed mesh on one side and a removable mesh on the other side. Adults were allowed to emerge for 5–6 days and were maintained on a 10% sugar solution for 12-36 hours prior to releases. We estimated that the releases were slightly male biased with an average female:male ratio within the devices of 3:4. The release devices were then stacked, sugar-free into boxes for transport to the release site.

### Wolbachia deployments

Mosquito deployments took place over a release area of 40 km^2^ during a period of 35 months (February 2017 - December 2019). Adult *w*Mel-infected mosquitoes were released weekly from a moving vehicle. In Zones 1 - 3 mosquito release points were initially determined using a 50 meter grid overlaid on the release areas, with one release point per grid square. In Zone 4 the density of release points was adjusted for the residential population in each neighbourhood, with the aim of releasing a cumulative total of 100 mosquitoes per resident (average distance between release points on a regular grid was 41 meters). In all areas, the initial release points determined on the grids were then distributed to the nearest vehicle-accessible road for vehicle releases (Figure S3). Releases were staged throughout the urban constructed areas in each release zone. Green non-constructed areas were excluded from releases as they provide less favourable habitats for *Ae. aegupti* and had few or no human residents. Initial release periods were 10-16 weeks duration, with subsequent re-releases conducted in local areas where *w*Mel prevalence was <40% in 3 consecutive monitoring events as measured at least 4 weeks after the conclusion of releases. This 40% threshold was based on previous estimates of the unstable equilibrium point for *w*Mel, above which invasion can occur.^25^ This resulted in re-releases being conducted in approximately 30% of the initial release areas. Most areas of Zones 1 and 2 had two periods of releases, Zone 3 had three periods of releases and Zone 4 only 1 release period.

### Wolbachia monitoring

Mosquitoes were collected weekly during and after releases using a network of BG Sentinel traps (Biogents AG, Regensburg, Germany, Product number NR10030) at an average density of 16 BG traps/km^2^ throughout release areas (Figure S4). Once *w*Mel prevalence was detected at >60% in 3 consecutive monitoring events measured at least 4 weeks after the conclusion of releases, trap numbers were reduced to 50% within a neighbourhood (Figure S4). Mosquitoes were sent to the laboratory for sorting, morphological identification and counting. The number of mosquitoes caught in each BG trap was recorded by species, sex, and in total. Mosquito samples were stored in 70% ethanol until screening for *w*Mel-strain *Wolbachia*. Screening was performed weekly until week ending 8 April 2018 and fortnightly thereafter.

### Wolbachia molecular detection

A maximum of 10 adult *Ae. aegypti* per BG trap per collection were screened for the presence of *w*Mel using either quantitative polymerase chain reaction (qPCR), or a colorimetric loop-mediated isothermal amplification (LAMP) assay. Taqman qPCR was performed on a Roche LightCycler 480 as described previously.^16,26^ Briefly, the qPCR cycling program consisted of a denaturation at 95°C for 5 min followed by 40 cycles of PCR (denaturation at 95 °C for 10 min, annealing at 60 °C for 30 sec, and extension at 72 °C for 1 sec with single acquisition) followed by a cooling down step at 40°C for 30 sec. LAMP reactions were performed in a Bio-Rad C1000 96-well PCR thermocycler with a 30min incubation at 65°C as previously described.^16^ Individual reactions consisted of 2X WarmStartR Colorimetric LAMP Master Mix (New England BioLabs, Cat# M1800S), primers and 1 μL of target DNA from a 50μl single mosquito squash buffer extraction assay, in a total reaction volume of 17 μL. An individual mosquito was scored as positive for *Wolbachia* if the Cp (crossing point) value in qPCR was below 28, or if the well in the LAMP assay was yellow upon visual inspection. Equivocal results were counted as negative. Details of primer and probe nucleotide sequences are included in the supplementary materials.

### Epidemiological data

Data on dengue and chikungunya cases notified to the Brazilian national disease surveillance system (SINAN) were used to evaluate the epidemiological impact of *Wolbachia* releases. Reporting of both diseases is mandatory in Brazil. Dengue notification data for Niterói is available from SINAN since 2007 and chikungunya since 2015. Notified dengue and chikungunya cases reported to SINAN are predominantly suspected cases based on a clinical case definition.^27^

Between 2007 - 2014, approximately 15% of notified dengue cases had supportive laboratory test results, usually from IgM serology. Since the Zika epidemic in Brazil in 2015, laboratory confirmation of dengue has relied on PCR only due to cross-reactive serological responses, and only one dengue case notified in 2015 - 2020 included laboratory confirmation. For chikungunya, 24% of cases notified in 2015 - 2020 had supportive IgM serology results. For the purpose of this analysis, we include all notified dengue and chikungunya cases (suspected and laboratory confirmed).

Anonymized disaggregate data on notified suspected and laboratory-confirmed dengue, severe dengue, chikungunya and Zika cases were obtained from the SINAN system through the Health Secretariat of Niterói, for the period from January 2007 (January 2015 for chikungunya and Zika) to June 2020. Population data by neighbourhood of residence from the Brazilian 2010 census (IBGE) was used to estimate the population in each *Wolbachia* release zone.

### Measurement of epidemiological impact

The *w*Mel intervention effect was estimated using controlled interrupted time series analysis performed separately for each release zone compared with the pre-defined control area, and for the aggregate release area compared with the control area, as described in a published study protocol.^21^ The primary analysis included data from January 2007 (dengue) or January 2015 (chikungunya and Zika), until June 2020, encompassing 8-37 months of post-intervention observations. For zone-level analyses, negative binomial regression was used to model monthly dengue, chikungunya and Zika case counts in the intervention and control areas, with an offset for population size. Seasonal variability in disease incidence was controlled using flexible cubic splines with knots placed at 6-monthly intervals. For the primary analysis, a binary ‘group’ variable indicated the study arm (intervention or control). A binary ‘treatment’ variable distinguished the pre-intervention period and the post-intervention period. The zone-level post-intervention period was defined as four weeks after *w*Mel releases had commenced throughout the whole zone; the corresponding post-intervention period was also applied to the control area for each zone-level analysis. The intervention effect was estimated from the interaction between the ‘group’ and ‘treatment’ variables, which allows explicitly for a level change in the outcome (dengue/chikungunya/Zika case incidence) in both intervention and control areas in the post-intervention period. Robust standard errors were used to account for autocorrelation and heteroskedasticity. A mixed-effects negative binomial regression was used to model monthly dengue, chikungunya or Zika case counts in the aggregate release area compared with the control area, with an offset for population size and controlling for seasonal variability in incidence using flexible cubic splines with knots placed at 6-monthly intervals. Clustering of dengue/chikungunya/Zika cases by release zone was modelled as a random effect by including a random intercept at the zone level and allowing for a random slope on the intervention. A binary ‘treatment’ variable distinguished the pre-intervention period and the post-intervention period, with the control area classified as ‘pre-intervention’ throughout. Robust standard errors were used to account for autocorrelation and heteroskedasticity. The zone-level and aggregate release area analyses included the pilot release area of Jurujuba within zone 1.

To account for within-zone heterogeneity in *w*Mel establishment and dengue incidence, a secondary neighbourhood-level analysis was also performed in which *Wolbachia* exposure was determined by the measured *w*Mel prevalence in *Ae. aegypti* collected from each neighbourhood, and a three-month moving average calculated to smooth the variability in monthly *w*Mel prevalence, categorised into quintiles of exposure. In zone 1 we excluded the neighbourhood of Jurujuba where pilot *w*Mel releases were staggered across seven sectors over a period of 16 months and *w*Mel monitoring was initially done only in small pockets of the neighbourhood where releases had already occurred, because the *w*Mel time-series during this staged release period was not representative of the whole of Jurujuba neighbourhood (whereas the dengue cases data was aggregate for the whole neighbourhood). This analysis included data to March 2020 only, as no *Wolbachia* monitoring was possible April – June 2020 due to restrictions on movement in response to the Covid-19 pandemic. Mixed-effects negative binomial regression was used to model monthly dengue case notifications by neighbourhood, in each of the four release zones individually and in all zones combined, compared with the pre-specified control zone. The model included population size as an offset and neighbourhood as a random effect. Given the large number of zero dengue case counts (zero-inflation) at the neighbourhood level, an alternative analysis using a zero-inflated negative-binomial model with robust standard errors to account for clustering was considered. Model fit was not improved by accounting for zero-inflation, as assessed using the Akaike Information Criterion (AIC), and was thus not used in the analyses. This secondary analysis was not performed for chikungunya or Zika due to the sparsity of case data at the neighbourhood level.

### Sensitivity analyses

As a sensitivity analysis, we excluded pre-intervention observations prior to 2012 to achieve greater balance between pre-intervention and post-intervention period lengths while maintaining sufficient data to inform on pre-intervention trends.^28^

### Power estimation

Power was estimated for the ITS analysis using 1000 simulated datasets drawn from a negative binomial distribution fitted to a ten-year time series (2007–2016) prior to *Wolbachia* deployment, of monthly dengue case notifications from release and control zones in Niterói and Rio de Janeiro. The simulated time series of dengue case numbers in the control zones as well as the pre-*Wolbachia* release dengue case numbers in the treated zones were drawn directly from this model-generated distribution. Post-*Wolbachia* release dengue case numbers in the treated zones were drawn from the same model-generated distribution, modified by an additional parameter for an intervention effect of Relative Risks = 0.6, 0.5, 0.4, 0.3. For each of these four ‘true’ effect sizes and a null effect (RR = 1), applied to each of the 1000 simulated time series, the ‘observed’ effect size was calculated from a negative binomial regression model of monthly case counts in the treated and untreated zones, as described above. Post-intervention time periods of 1, 2 or 3 years were simulated, with the pre-intervention period fixed at 7 years. The estimated power to detect a given effect size was determined as the proportion of the 1000 simulated scenarios in which a significant intervention effect (p<0.05) was observed. These simulations indicate 80% power to detect a reduction in dengue incidence of 50% or greater after three years of post-intervention observations, and a reduction of 60% or greater after two years.

## Results

### Wolbachia establishment in Niterói

Awareness (prior knowledge of the *Wolbachia* method) ranged from 36 to 50% and acceptance (agreement with the proposed *w*Mel releases in the neighbourhood) ranged from 65 to 92%, in the public survey conducted prior to releases in Niterói. No negative media nor negative community incidents were registered, and the Community Reference Group endorsed the start of releases.

Heterogeneity in *w*Mel *Wolbachia* establishment was observed in three of the four release zones (Figure 2). In the initial release area of zone 1, *Wolbachia* prevalence was greater than 80% in the first quarter of 2020 (up to 11 months post-release) and there was low variability across the neighbourhoods. Local *w*Mel introgression has been more variable in zones 2 and 3, with a median *w*Mel prevalence of 40 -70% among neighbourhoods during the post-release period (11 months and 9 months, respectively). In zone 4, a longer post-intervention observation period is required to evaluate the trajectory of *w*Mel establishment. *Aedes albopictus* is present throughout the city and was detected at a similar abundance in our monitoring network during and after releases (Figure S5).

**Figure 2:**
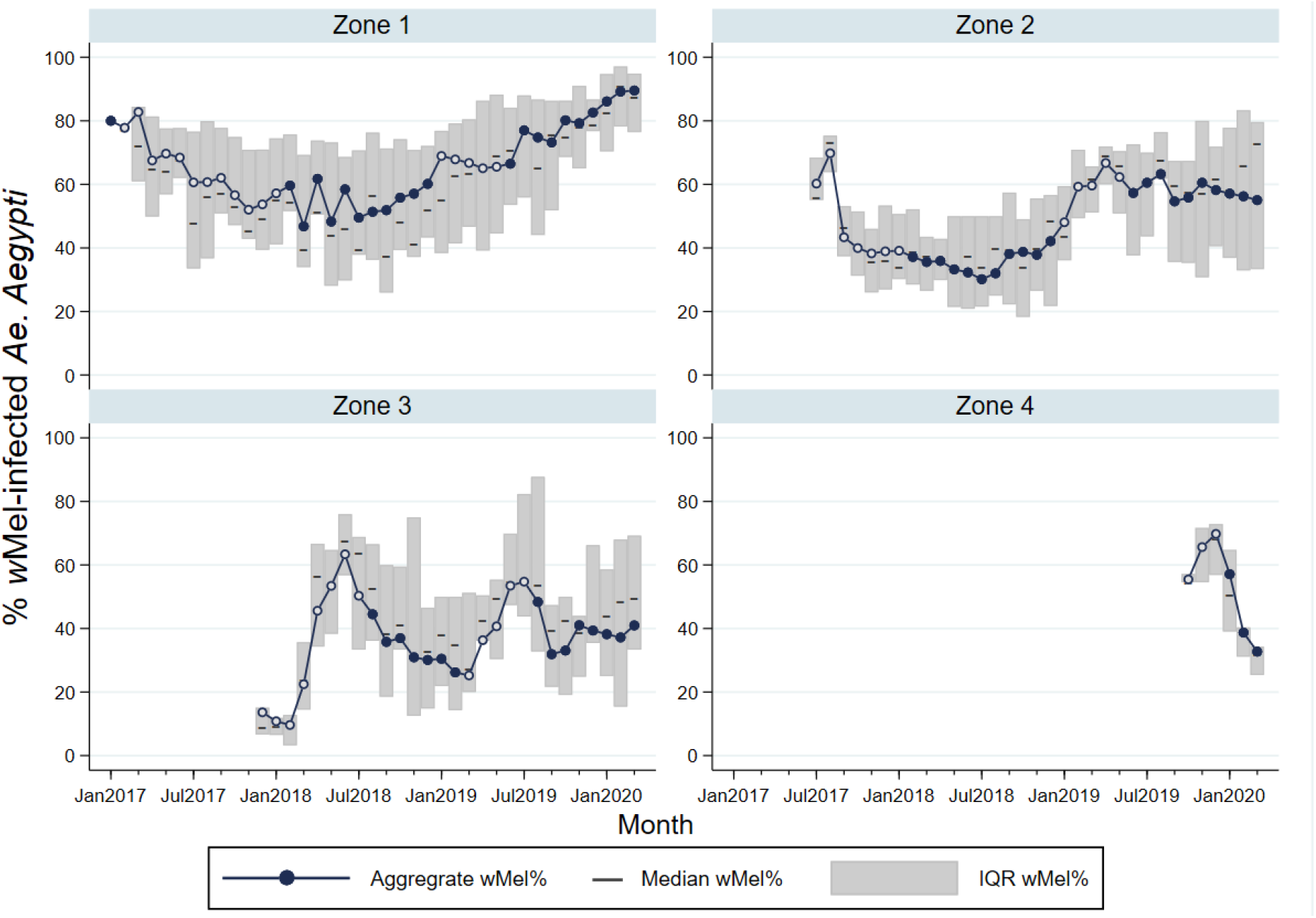
*w*Mel infection prevalence in *Aedes aegypti* mosquitoes collected from each release zone, during and after releases. Circle markers represent the aggregate *w*Mel infection prevalence in each zone in each calendar month from January 2017 to March 2020. Open circles indicate months when *Wolbachia* releases took place in any part of that zone; filled circles are months with no releases. Horizontal lines represent the median *w*Mel infection rate among the individual neighbourhoods in each zone (n=4 neighbourhoods in Zone 1; n=11 in Zone 2; n=13 in Zone 3; n=5 in Zone 4). Shaded bars show the interquartile range (IQR) of *w*Mel infection rates among the individual neighbourhoods in each zone, each month. Note that in January and February 2017, the only BG traps in Zone 1 were in the Jurujuba pilot release area where releases and monitoring had been ongoing throughout 2015-2016;^20^ BG monitoring in March-April 2017 had commenced in 2/4 neighbourhoods and from May 2017 in all 4 Zone 1 neighbourhoods. In Zone 2, BG monitoring in July-Sept 2017 had commenced in 7/11 neighbourhoods and from Oct 2017 in all 11 neighbourhoods. In Zone 3 and Zone 4, BG monitoring commenced in all neighbourhoods from Dec 2017 and Oct 2019, respectively.

### *Arboviral disease trends pre- and post-*Wolbachia *intervention*

During the ten years prior to the start of scaled *Wolbachia* mosquito releases in Niterói in early 2017, seasonal peaks in dengue case notifications occurred each year (Figure 3A), usually in March and April (Figure 3B). A median of 2,818 dengue cases were notified each year 2007 - 2016 (per capita incidence 581/100,000 population), with a minimum of 366 cases in 2014 (75/100,000) following a maximum of 11,618 in 2013 (2,396/100,000). In the three years following the start of phased *Wolbachia* releases, annual city-wide dengue case notifications were 895, 1,729 and 378 in 2017, 2018 and 2019 respectively, and the seasonal peaks in dengue incidence occurred predominantly in the areas of Niterói that had not yet received *Wolbachia* deployments (Figure 4).

**Figure 3:**
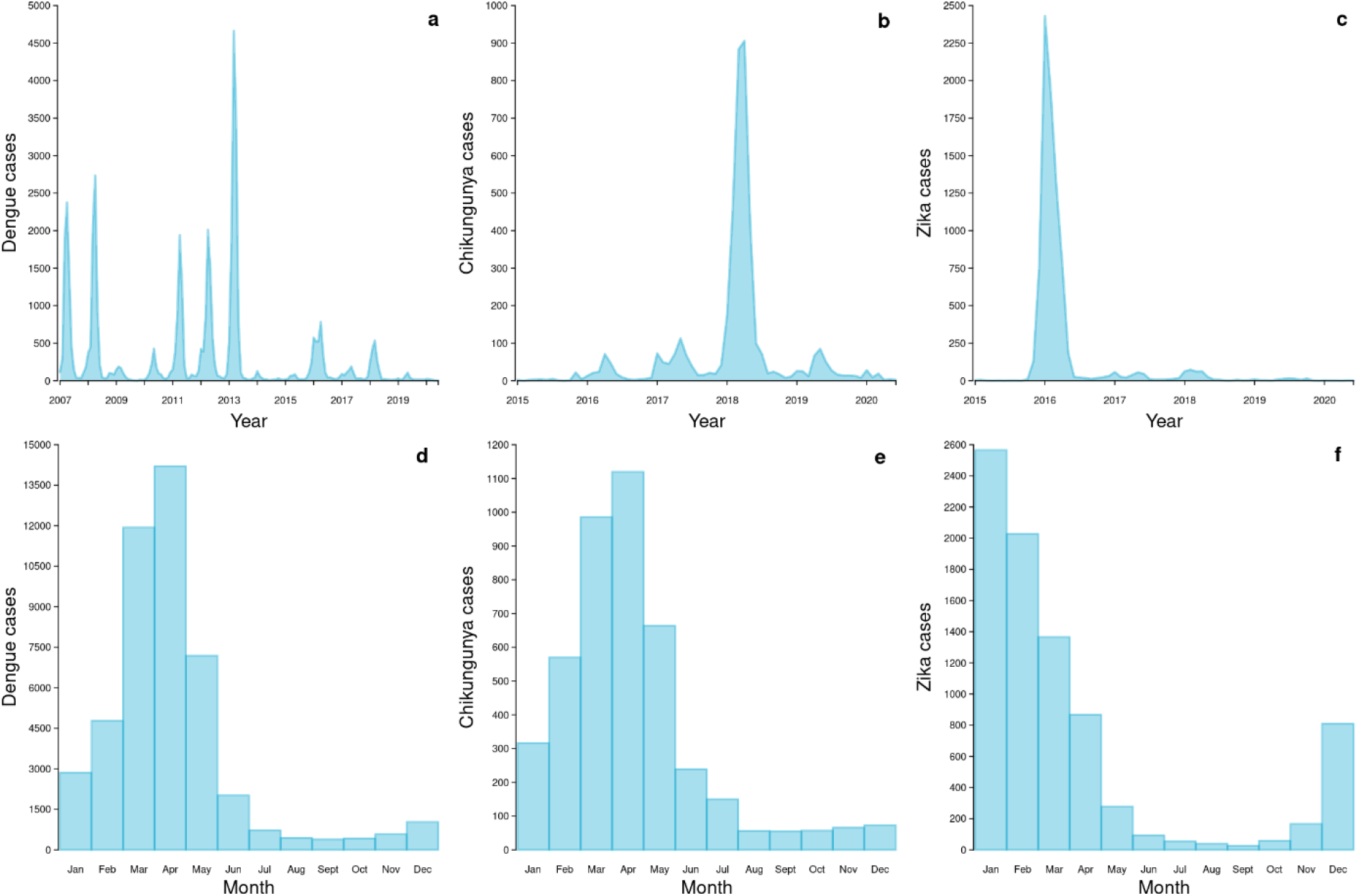
Dengue, chikungunya and Zika time series and seasonality in Niterói. Monthly dengue (a), chikungunya (b) and Zika (c) case notifications in Niterói from January 2007 (dengue) or January 2015 (chikungunya/Zika) to June 2020, and dengue (d), chikungunya (e) and Zika (f) case notifications aggregated by calendar month, across the same period.

**Figure 4:**
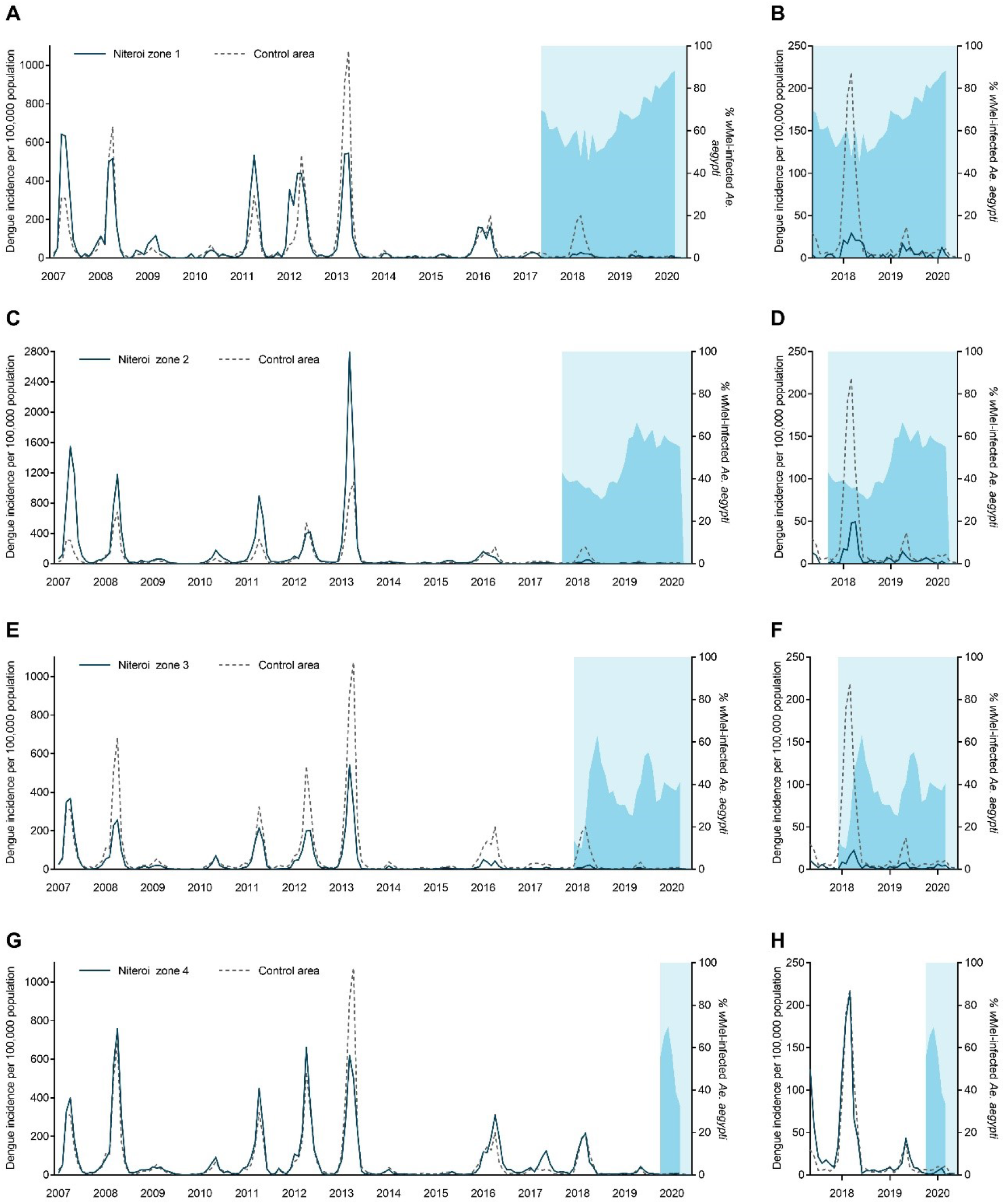
Dengue incidence and *w*Mel infection prevalence in local *Aedes aegypti* mosquito populations, by release zone. **Panels A**,**C**,**E**,**G:** Lines show the monthly incidence of dengue case notifications per 100,000 population (left-hand Y axis) in Niterói release zones 1 - 4 (solid line in each panel) compared with the untreated control zone (dashed line), January 2007 - June 2020. Light blue shading indicates the beginning of the epidemiological monitoring period in each zone, one month after initial releases were completed in each respective zone. Darker blue shading indicates the aggregate *w*Mel infection prevalence (right-hand Y axis) in each zone in each calendar month from the start of the epidemiological monitoring period until March 2020 (no *w*Mel monitoring April - June 2020). **Panels B**,**D**,**F**,**H** show the same data but zoomed into the period from May 2017 – March 2020 and with the dengue incidence axis rescaled, to show more clearly the trends in release and control zones in the post-intervention period.

Chikungunya surveillance commenced in January 2015. Between 44 and 533 chikungunya cases were notified annually in Niterói in 2015 - 2019, with the exception of 2018 when an explosive outbreak resulted in 3091 reported cases; 95% of those occurred in the six months January to June. The highest per capita incidence of chikungunya during the 2018 outbreak was in the untreated control zone (1,413 cases/100,000 population; Figure 5), followed by Zone 4 where *Wolbachia* deployments had not yet commenced (958/100,000). In Zones 1, 2, and 3 where deployments were underway and zone-level *Wolbachia* prevalence was between 20 – 55%, the incidence of chikungunya case notifications during the 2018 outbreak was 106/100,000, 244/100,000 and 201/100,000, respectively.

**Figure 5:**
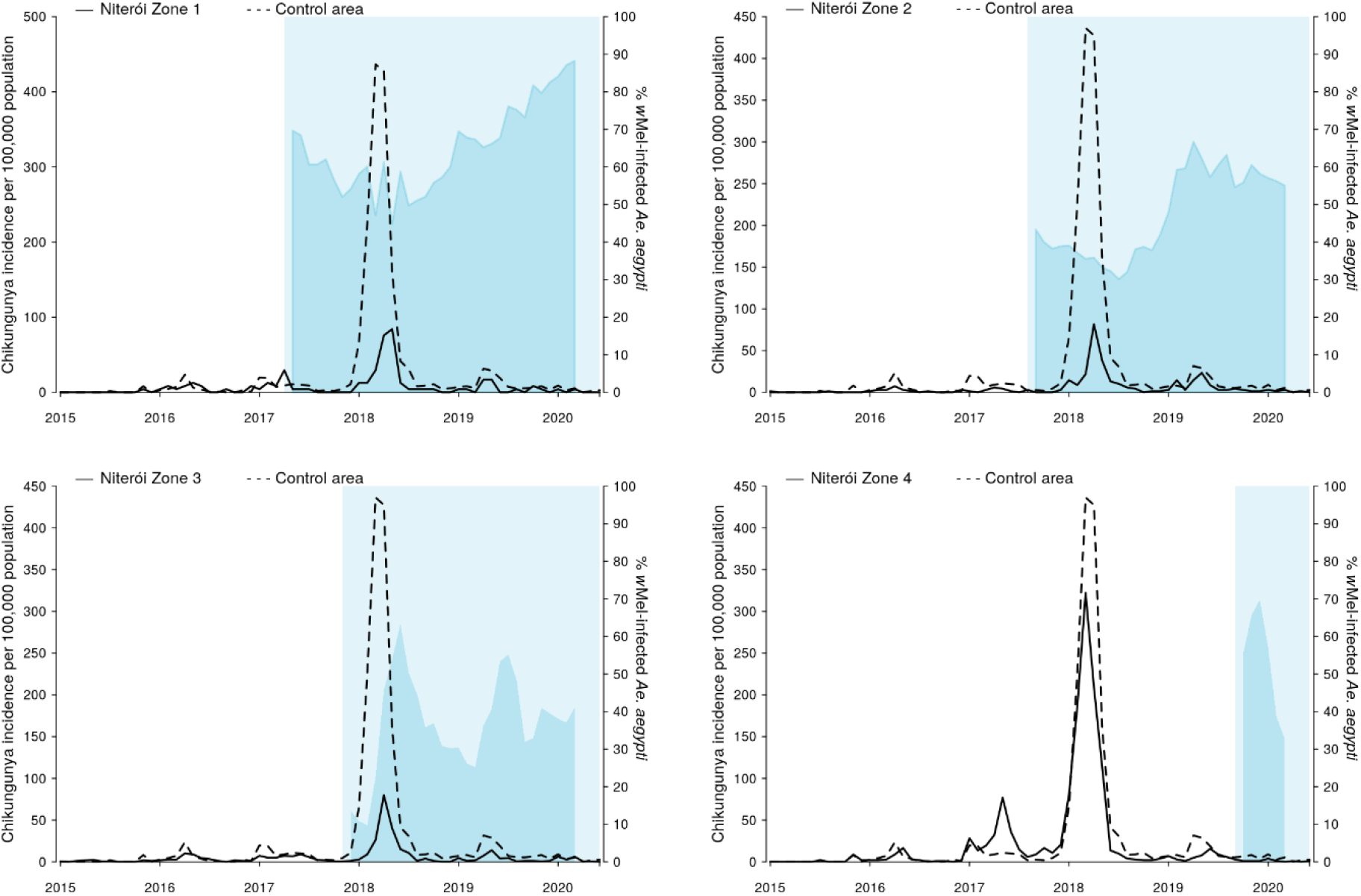
Chikungunya incidence and *w*Mel infection prevalence in local *Aedes aegypti* mosquito populations, by release zone. Lines show the monthly incidence of chikungunya case notifications per 100,000 population (left-hand Y axis) in Niterói release zones 1 - 4 (solid line in each panel) compared with the untreated control zone (dashed line), January 2015 - June 2020. Light blue shading indicates the beginning of the epidemiological monitoring period in each zone, one month after initial releases were completed in each respective zone. Darker blue shading indicates the aggregate *w*Mel infection prevalence (right-hand Y axis) in each zone in each calendar month from the start of the epidemiological monitoring period until March 2020 (no *w*Mel monitoring April - June 2020).

There were 8,247 Zika cases reported in Niterói between 2015 and June 2020, 91% (n=7,532) of which were reported in 2015-2016 when Brazil experienced an unprecedented Zika outbreak (Figure 6). From 2017, when phased *w*Mel deployments began in Niterói, until June 2020 a total of 715 Zika cases were notified in Niterói, of which of 95 were reported from areas where *w*Mel deployments had already occurred: 12 in zone 1, 28 in zone 2, 48 in zone 3, and 7 in zone 4.

**Figure 6:**
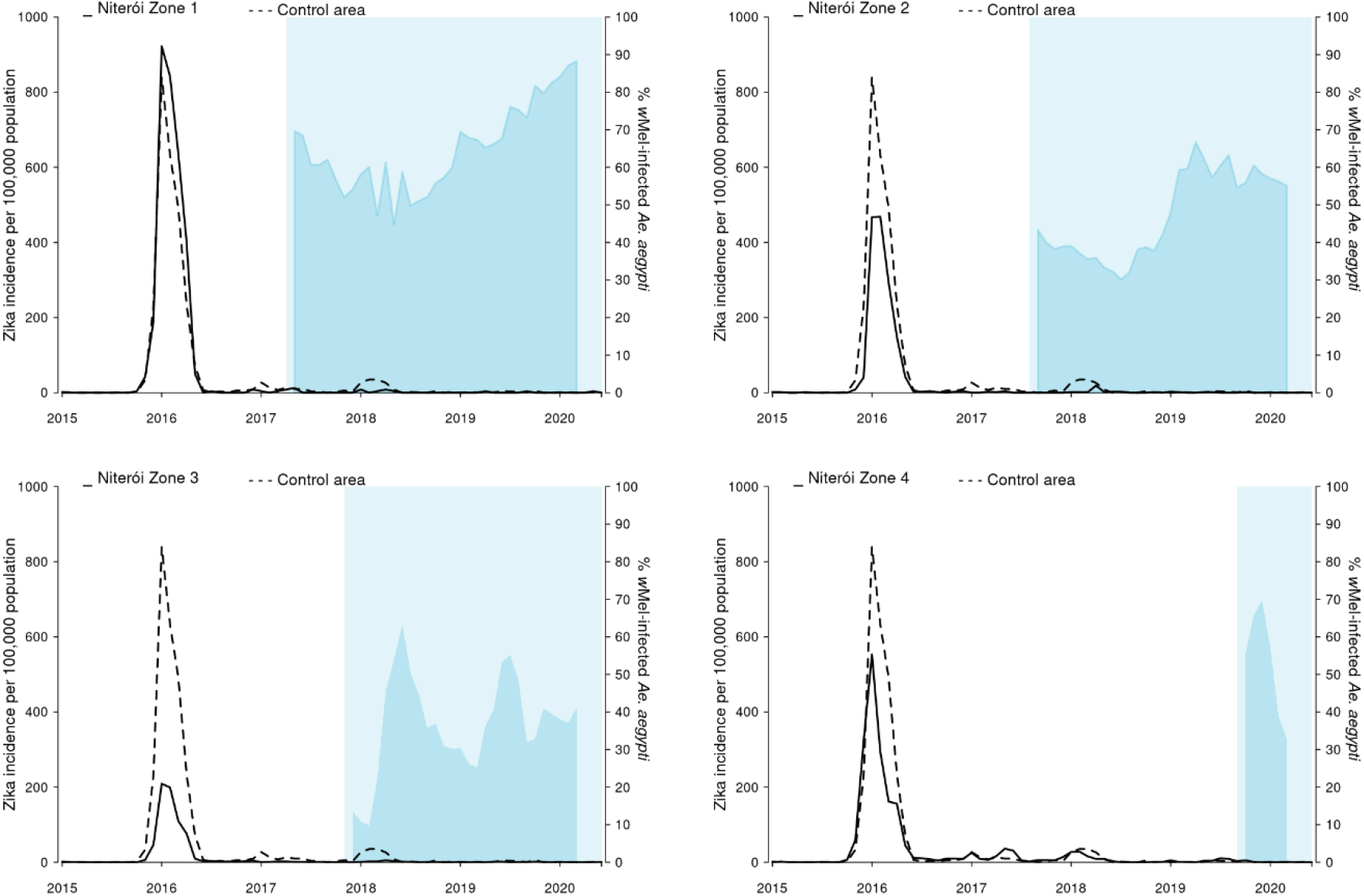
Zika incidence and *w*Mel infection prevalence in local *Aedes aegypti* mosquito populations, by release zone. Lines show the monthly incidence of Zika case notifications per 100,000 population (left-hand Y axis) in Niterói release zones 1 - 4 (solid line in each panel) compared with the untreated control zone (dashed line), January 2015 - June 2020. Light blue shading indicates the beginning of the epidemiological monitoring period in each zone, one month after initial releases were completed in each respective zone. Darker blue shading indicates the aggregate *w*Mel infection prevalence (right-hand Y axis) in each zone in each calendar month from the start of the epidemiological monitoring period until March 2020 (no *w*Mel monitoring April - June 2020).

### *Reduction in dengue, chikungunya and Zika incidence post-*Wolbachia *intervention*

Using interrupted time series (ITS) analysis to account for underlying temporal trends in case incidence and staggered implementation of the intervention, we found that *w*Mel *Wolbachia* deployments were associated with a significant reduction in dengue incidence in each of the four release zones (Figure 6a). The magnitude of this reduction ranged from 46.0% (95%CI 21.0, 63.0) in Zone 3 to 75.9% (95%CI 62.1, 84.7) in Zone 2. Overall, *Wolbachia* deployments were associated with a 69.4% (95%CI 54.4, 79.4) reduction in dengue incidence in Niterói (Figure 7A; Table S1).

**Figure 7:**
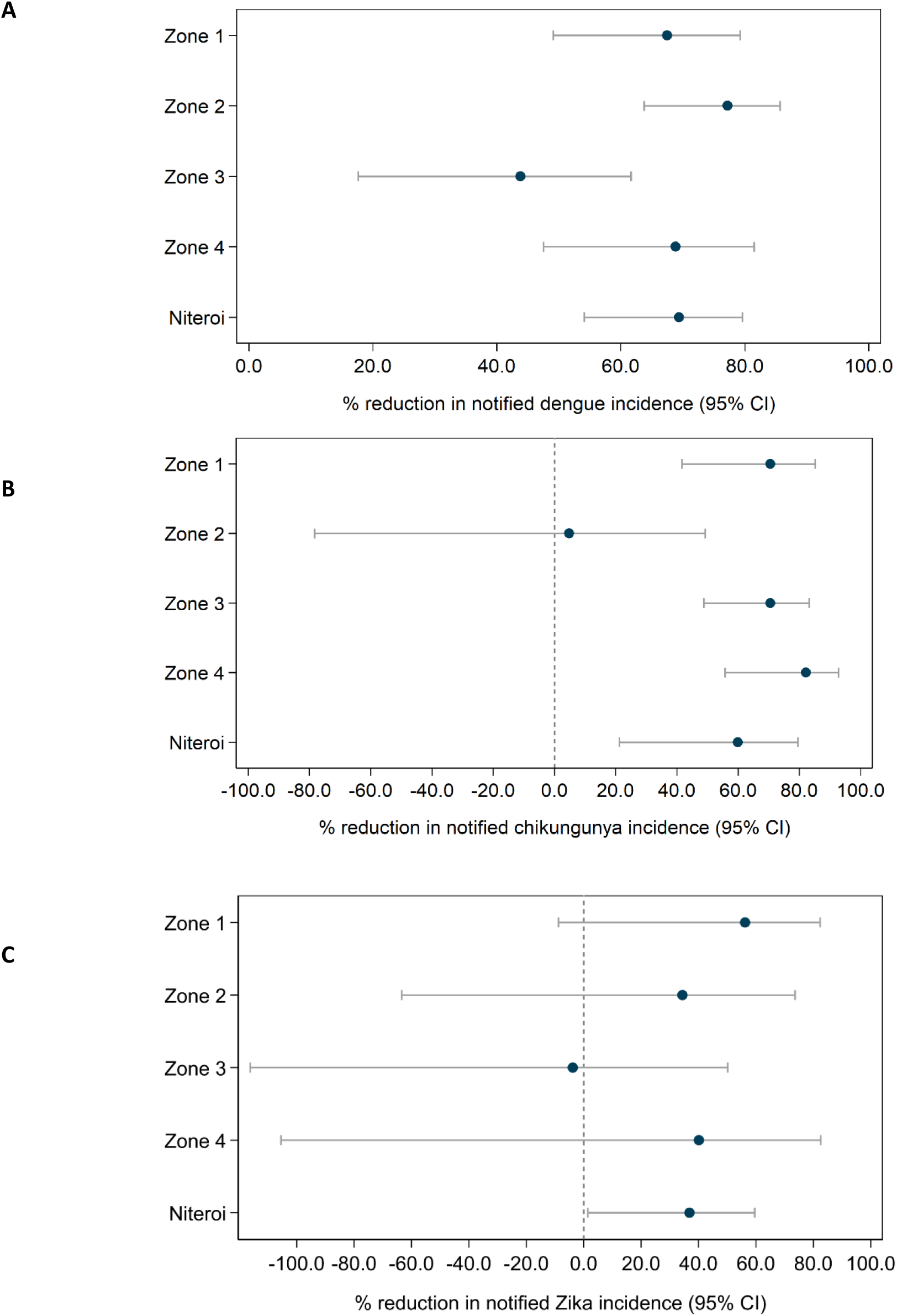
Estimated reduction in the incidence of dengue (A) chikungunya (B) and Zika (C) following *Wolbachia* deployments in Niterói, in each release zone individually and in the aggregate release area. Point estimates (circles) and 95% confidence intervals (horizontal bars) from controlled interrupted time series analysis of monthly dengue (Jan 2007 – June 2020), chikungunya and Zika (Jan 2015 – June 2020) case notifications to the Brazilian national disease surveillance system.

This *Wolbachia* intervention effect against dengue was also apparent overall, and in each zone, in the neighbourhood-level analysis that considered quintiles of *w*Mel prevalence in local *Ae. aegypti* populations, although we found evidence of only marginal additional reductions in dengue incidence at higher levels of *Wolbachia* beyond 20 – 40% *w*Mel prevalence (Figure S6; Table S2). There was substantial month-to-month variation in *w*Mel quintiles within neighbourhoods (Figure S7), which was reduced but not removed by taking a three-month moving average of *w*Mel prevalence. The result were little changed in the sensitivity analysis, which excluded pre-intervention observations prior to 2012 (Figure S8; Table S3).

A total of 897 severe dengue cases were reported in Niterói between 2007 and early 2020, 691 of which were from one of the four intervention zones and 206 from the control zone. Only three of these cases occurred in the post-intervention period, two in zone 2 and one in zone 3. The control zone has not had any severe dengue cases reported since 2016. These numbers were too sparse to be analysed using our ITS model, even when allowing for zero-inflation.

We found in ITS analysis that chikungunya incidence was also significantly reduced following *Wolbachia* deployments in Niterói as a whole (56.3% reduction in incidence; 95%CI 15.9, 77.3) and in three of the four individual release zones (Figure 7B; Table S1). Zika incidence was reduced by 37% (95%CI 1.5, 59.5) following *Wolbachia* deployments in Niterói as a whole, though not in individual release zones (Figure 7C; Table S1).

## Discussion

Large-scale phased deployments of *w*Mel strain *Wolbachia*-infected *Aedes aegypti* mosquitoes in Niterói, Brazil during 2017 - 2019, resulted in *w*Mel establishment in local *Ae. aegypti* populations at an infection frequency of 33 – 90% by March 2020, when field monitoring was paused due to the emergence of SARS-CoV-2 in Brazil. More than one-quarter of the total 373,000 residents of the intervention area were living in neighbourhoods where local *w*Mel prevalence was 60% or greater by March 2020, predominantly in zones 1 and 2 where releases commenced earliest. In the remaining intervention areas, *w*Mel prevalence was more heterogeneous and a resumption of entomological monitoring is planned in order to evaluate the long-term trajectory of *w*Mel introgression into the local *Ae. aegypti* population.

Despite this heterogeneity in *Wolbachia* establishment, a significant reduction in the incidence of dengue, chikungunya and Zika case notifications was observed in *Wolbachia*-treated areas of Niterói, compared with a pre-defined untreated control area. This epidemiological impact on dengue was replicated across all four release zones, and in three of the four zones for chikungunya. Aggregate across the whole intervention area, the *w*Mel deployments were associated with a 69% reduction in dengue incidence, a 56% reduction in chikungunya incidence and a 37% reduction in Zika incidence. Given the recognised lack of evidence for efficacy of routinely available approaches to arboviral disease control^4^ based on elimination of breeding sites and insecticide-based suppression of adult mosquito populations, and considering the magnitude of the historical burden of *Aedes*-borne disease in Niterói, an intervention effect of this magnitude represents a substantial public health benefit.

Results from a recent cluster randomised trial of *w*Mel-infected *Ae aegypti* deployments in Yogyakarta Indonesia demonstrated 77% efficacy in preventing virologically confirmed dengue cases,^18^ with comparable efficacy against all four dengue virus serotypes. Previous non-randomised controlled field trials in Indonesia^15^ and northern Australia^16,17^ demonstrated 76% and 96% effectiveness, respectively, in reducing the incidence of dengue cases notified to routine disease surveillance systems. In each of those sites the trajectory of *w*Mel establishment was more rapid and more homogeneous across the release area than observed in Niterói. In the present study, the epidemiological impact in the area of Niterói where *w*Mel introgression occurred most rapidly and homogeneously (Zone 1) was highly comparable with the Indonesian studies: 77% (95%CI 64, 86). Another *Wolbachia* strain, *w*AlbB, has been successful introgressed into *Ae. aegypti* field populations in Kuala Lumpur, Malaysia,^29^ although with instability in *w*AlbB frequencies in some release areas after cessation of releases, which the authors attributed to immigration of wild-type mosquitoes into the small release sites (area 0.05 – 0.73 km^2^) from surrounding untreated areas.

The reasons for slower and more heterogeneous *w*Mel introgression here, compared to Indonesia and Australia, are not fully understood. A likely contributing factor is that these scaled deployments have largely used adult mosquitoes released from vehicles, which does not deliver as spatially homogeneous a deployment as occurred previously in Indonesia and Australia. In contrast, the small-scale pilot releases in the Jurujuba neighbourhood of Niterói in 2015 achieved rapid and sustained introgression of *w*Mel after 8 – 31 weeks of egg-based releases.^20^ Additionally, Niterói release areas were complex urban environments with high rise areas and large informal settlements, where field activities were frequently interrupted by security issues and where physical barriers to spread,^30^ spatial heterogeneity in mosquito abundance,^31^ and limited mosquito dispersal^32^ could have contributed to slower *w*Mel introgression. The wild-type egg bank in such a setting is likely to be large and spatially heterogeneous, and would take time to be depleted, which is likely to have contributed to the heterogeneity in *w*Mel introgression and intermediate frequencies of *Wolbachia* observed in our study. Regular monitoring of the *w*Mel-*Ae. aegypti* broodstock has demonstrated insecticide susceptibility profiles comparable with wild-type material, so a concern of increased susceptibility to insecticide is not considered to be an issue here. Impaired maternal transmission by *w*Mel -infected females^33,34^ and loss of induction of cytoplasmic incompatibility by *w*Mel -infected males^35^ has been observed by others at high, but field relevant, temperatures. Exposure of immature *Ae. aegypti* to very high temperatures in small water containers cannot be excluded as a contributing factor to the *w*Mel introgression patterns observed in Niteroi, especially in the more informal settlements where the urban landscape is more vulnerable to temperature variations. Entomological monitoring in future years will help clarify the long-term trajectory of *w*Mel introgression in Niterói.

In large and complex urban environments, a homogeneous high level of introgression of *w*Mel may prove operationally challenging and slow to achieve, even with optimised release methods and longer post-release monitoring. This poses the question of what minimum threshold of *w*Mel prevalence is needed to achieve interruption of local arbovirus transmission, and whether a dose-response relationship is observed between *w*Mel prevalence and disease reduction. Predictions from mathematical models have suggested that even in conservative scenarios where scaled *Wolbachia* deployments only reduce the reproduction number (R0) of dengue by 50%, this could lead to reductions in global case incidence of 70%,^36^ although the impact is predicted to be highly spatially heterogeneous, with smaller relative reductions in areas with highest transmission intensity. Our findings support this prediction of epidemiological impact with imperfect *w*Mel-mediated transmission blocking, by demonstrating that measurable reductions in dengue, chikungunya and Zika disease accrue even at a moderate prevalence of *w*Mel in local *Ae. aegypti* populations. A secondary analysis based on measured *w*Mel prevalence and dengue case notifications at the neighbourhood-level found only a marginal increase in the *w*Mel intervention effect beyond 20 – 40% prevalence, which was unexpected. This analysis also indicated substantial variability in *w*Mel prevalence over time (within neighbourhoods). This may be attributable in part to sampling variability due to small *Ae. aegypti* catch numbers in some areas but may also indicate true local instability in *Wolbachia* levels. When combined with people’s mobility and risk of acquiring dengue outside their neighbourhood of residence these factors may help explain the non-linear association between measured neighbourhood-level monthly *w*Mel infection prevalence and dengue risk. The absolute abundance of wild-type *Ae. aegypti*, independent of *w*Mel prevalence, is also relevant to understanding local dengue risk and could not be accounted for in our time series analyses because of a lack of baseline (pre-intervention) mosquito collection data. We cannot exclude that incompatibility within the population of *w*Mel positive and negative *Ae. aegypti* could impact overall population size and contribute to the observed epidemiological outcomes. Overall, a contribution of indirect effects, confounding by mosquito population size, and imperfect *w*Mel exposure measurement to the observation of an epidemiological impact even at moderate *w*Mel prevalence cannot be excluded, and this observation needs replication in other settings.

This study has some limitations. Deployments of *w*Mel-infected *Ae. aegypti* were not randomised, so there is the potential for measurement of the intervention effect to be confounded by other factors that differ between the release areas and the pre-defined control area. Routine disease surveillance data is imperfect both in specificity (not all notified cases are true dengue/chikungunya/ZIka cases) and in sensitivity (not all dengue/chikungunya/Zika cases are notified). However, the risk of these factors influencing the measurement of the epidemiological endpoint here is reduced by the inclusion of a parallel control with a historical dengue time series that is highly synchronous with each of the release areas for ten years pre-intervention. The replication of the dengue intervention effect in each of the four release zones, and for chikungunya in three zones, also mitigates the possibility that any parallel change in vector control practices or healthcare seeking behaviour in intervention areas could have confounded the observed result. For chikungunya and Zika, there is substantial uncertainty around the point estimate of the intervention effect because case notifications for these two diseases were very sparse in both release and control areas outside of a single large outbreak in 2018 and 2015-16, We have demonstrated that *w*Mel introgression can be achieved across a large and complex urban environment over a period of three years to a prevalence in local *Ae. aegypti* which, while still heterogeneous, is sufficient to result in a measurable reduction in dengue, chikungunya and Zika case incidence. Ongoing entomological and epidemiological monitoring will provide additional information on the trajectory of *w*Mel establishment in areas where releases have occurred more recently, or introgression has been slower, and on the full magnitude and durability of the public health benefit.

## Data Availability

The data underlying the results presented in this study are publicly available at 10.6084/m9.figshare.13662203.v3 (zone-level data) and 10.6084/m9.figshare.13662230.v2 (neighbourhood-level data).

## Acknowledgments

The authors acknowledge the municipality of Niterói for their partnership and logistical support for this study.

## References

1. Cattarino L, Rodriguez-Barraquer I, Imai N, Cummings DAT, Ferguson NM. Mapping global variation in dengue transmission intensity. Sci Transl Med 2020;12.

2. Stanaway JD, Shepard DS, Undurraga EA, et al. The global burden of dengue: an analysis from the Global Burden of Disease Study 2013. Lancet Infect Dis 2016;16:712–23.

3. Shepard DS, Undurraga EA, Halasa YA, Stanaway JD. The global economic burden of dengue: a systematic analysis. Lancet Infect Dis 2016;16:935–41.

4. Bowman LR, Donegan S, McCall PJ. Is dengue vector control deficient in effectiveness or evidence?: systematic review and meta-analysis. PLoS Negl Trop Dis 2016;10:e0004551.

5. Sim S, Ng LC, Lindsay SW, Wilson AL. A greener vision for vector control: The example of the Singapore dengue control programme. PLoS Negl Trop Dis 2020;14:e0008428.

6. Wilson AL, Boelaert M, Kleinschmidt I, et al. Evidence-based vector control? Improving the quality of vector control trials. Trends Parasitol 2015;31:380–90.

7. Aliota MT, Peinado SA, Velez ID, Osorio JE. The wMel strain of Wolbachia reduces transmission of Zika virus by Aedes aegypti. Sci Rep 2016;6:28792.

8. Aliota MT, Walker EC, Uribe Yepes A, Velez ID, Christensen BM, Osorio JE. The wMel strain of Wolbachia reduces transmission of chikungunya virus in Aedes aegypti. PLoS Negl Trop Dis 2016;10:e0004677.

9. Dutra HL, Rocha MN, Dias FB, Mansur SB, Caragata EP, Moreira LA. Wolbachia blocks currently circulating Zika virus isolates in Brazilian Aedes aegypti mosquitoes. Cell Host Microbe 2016;19:771–4.

10. Moreira LA, Iturbe-Ormaetxe I, Jeffery JA, et al. A Wolbachia symbiont in Aedes aegypti limits infection with dengue, chikungunya, and Plasmodium. Cell 2009;139:1268–78.

11. Pereira TN, Rocha MN, Sucupira PHF, Carvalho FD, Moreira LA. Wolbachia significantly impacts the vector competence of Aedes aegypti for Mayaro virus. Sci Rep 2018;8:6889.

12. van den Hurk AF, Hall-Mendelin S, Pyke AT, et al. Impact of Wolbachia on infection with chikungunya and yellow fever viruses in the mosquito vector Aedes aegypti. PLoS Negl Trop Dis 2012;6:e1892.

13. Walker T, Johnson PH, Moreira LA, et al. The wMel Wolbachia strain blocks dengue and invades caged Aedes aegypti populations. Nature 2011;476:450–3.

14. Ye YH, Carrasco AM, Frentiu FD, et al. Wolbachia reduces the transmission potential of dengue-infected Aedes aegypti. PLoS Negl Trop Dis 2015;9:e0003894.

15. Indriani C, Tantowijoyo W, Rances E, et al. Reduced dengue incidence following deployments of Wolbachia-infected Aedes aegypti in Yogyakarta, Indonesia: a quasi-experimental trial using controlled interrupted time series analysis. Gates Open Res 2020;4:50.

16. O’Neill SL, Ryan PA, Turley AP, et al. Scaled deployment of Wolbachia to protect the community from dengue and other Aedes transmitted arboviruses. Gates Open Res 2018;2:36.

17. Ryan PA, Turley AP, Wilson G, et al. Establishment of wMel Wolbachia in Aedes aegypti mosquitoes and reduction of local dengue transmission in Cairns and surrounding locations in northern Queensland, Australia. Gates Open Res 2019;3:1547.

18. Utarini A, Indriani C, Ahmad RA, et al. Efficacy of Wolbachia-infected mosquito deployments for dengue control. N Engl J Med 2021;In Press.

19. Garcia GA, Sylvestre G, Aguiar R, et al. Matching the genetics of released and local Aedes aegypti populations is critical to assure Wolbachia invasion. PLoS Negl Trop Dis 2019;13:e0007023.

20. Gesto JSM, Ribeiro GS, Rocha MN, et al. Reduced competence to arboviruses following the sustainable invasion of Wolbachia into native Aedes aegypti from Southeastern Brazil. Sci Rep 2021;11:10039.

21. Durovni B, Saraceni V, Eppinghaus A, et al. The impact of large-scale deployment of Wolbachia mosquitoes on dengue and other Aedes-borne diseases in Rio de Janeiro and Niteroi, Brazil: study protocol for a controlled interrupted time series analysis using routine disease surveillance data. F1000Res 2019;8:1328.

22. Costa GB, Smithyman R, O’Neill SL, Moreira LA. How to engage communities on a large scale? Lessons from World Mosquito Program in Rio de Janeiro, Brazil [version 1; peer review: 1 approved, 2 approved with reservations]. Gates Open Res 2020;4.

23. Garcia GA, Hoffmann AA, Maciel-de-Freitas R, Villela DAM. Aedes aegypti insecticide resistance underlies the success (and failure) of Wolbachia population replacement. Sci Rep 2020;10:63.

24. Rocha MN, Duarte MM, Mansur SB, et al. Pluripotency of Wolbachia against Arboviruses: the case of yellow fever. Gates Open Res 2019;3:161.

25. Hoffmann AA, Montgomery BL, Popovici J, et al. Successful establishment of Wolbachia in Aedes populations to suppress dengue transmission. Nature 2011;476:454–7.

26. Dar M, Giesler T, Richardson R, et al. Development of a novel ozone- and photo-stable HyPer5 red fluorescent dye for array CGH and microarray gene expression analysis with consistent performance irrespective of environmental conditions. BMC Biotechnol 2008;8:86.

27. Ministry of Health Brazil (Health Surveillance Secretariat). Guia de Vigilância em Saúde : volume único.

28. Bernal JL, Cummins S, Gasparrini A. Interrupted time series regression for the evaluation of public health interventions: a tutorial. Int J Epidemiol 2017;46:348–55.

29. Nazni WA, Hoffmann AA, NoorAfizah A, et al. Establishment of Wolbachia strain wAlbB in Malaysian populations of Aedes aegypti for dengue control. Curr Biol 2019;29:4241–8 e5.

30. Schmidt TL, Filipovic I, Hoffmann AA, Rasic G. Fine-scale landscape genomics helps explain the slow spatial spread of Wolbachia through the Aedes aegypti population in Cairns, Australia. Heredity (Edinb) 2018;120:386–95.

31. Hancock PA, Ritchie SA, Koenraadt CJM, Scott TW, Hoffmann AA, Godfray HCJ. Predicting the spatial dynamics of Wolbachia infections in Aedes aegypti arbovirus vector populations in heterogeneous landscapes. J Appl Ecol 2019;56:1674–86.

32. Jasper M, Schmidt TL, Ahmad NW, Sinkins SP, Hoffmann AA. A genomic approach to inferring kinship reveals limited intergenerational dispersal in the yellow fever mosquito. Mol Ecol Resour 2019;19:1254–64.

33. Mancini MV, Ant TH, Herd CS, et al. High temperature cycles result in maternal transmission and dengue infection differences between Wolbachia strains in Aedes aegypti. bioRxiv 2020.

34. Ross PA, Wiwatanaratanabutr I, Axford JK, White VL, Endersby-Harshman NM, Hoffmann AA. Wolbachia infections in Aedes aegypti differ markedly in their response to cyclical heat stress. PLoS Pathog 2017;13:e1006006.

35. Ross PA, Ritchie SA, Axford JK, Hoffmann AA. Loss of cytoplasmic incompatibility in Wolbachia-infected Aedes aegypti under field conditions. PLoS Negl Trop Dis 2019;13:e0007357.

36. Cattarino L, Rodriguez-Barraquer I, Imai N, Cummings DAT, Ferguson NM. Mapping global variation in dengue transmission intensity. Sci Transl Med 2020;12.

