## Supplementary material for "Effectiveness of *Wolbachia*-infected mosquito deployments in reducing the incidence of dengue and other *Aedes*-borne diseases in Niterói, Brazil: a quasi-experimental study"

**Supplementary Methods**:

**kdr genotyping**

Adult *Aedes aegypti* were genotyped by qPCR to detect single nucleotide polymorphisms (SNPs) at the 1016 (Val+ or Ilekdr) and 1534 (Phe+ or Cyskdr) positions of the voltage gated sodium channel gene (NaV), as previously reported.^1,2^ Amplification reaction was performed with LightCycler 480 Probes Master mix (Roche), 10ng of individual genomic DNA, and a set of primers and probes to detect kdr alleles (below) customized by Thermo Fisher Inc. under ID codes: AHS1DL6 (Val+1016Ilekdr) and AHUADFA (Phe+1534Cys). Thermal cycling was carried out on a Light Cycler 480 Instrument II (Roche), set to the following conditions: 95 ºC for 10 min (initial denaturation), and N cycles of 95 ºC for 15 s and 60 ºC for 30 s (single acquisition). N was set to 30, for amplifying Val+1016Ilekdr, or to 40, for Phe+1534Cyskdr. For each collection date, up to 90 samples were individually genotyped.

**Nucleotide sequences of primers and probes.** List of primers and probes used for the molecular diagnostics of *Wolbachia* by qPCR and LAMP, and for kdr genotyping.

| PRIMER | NUCLEOTIDE SEQUENCE (5’ à 3’) |
| --- | --- |
| Ae. aegypti RPS17 – qPCR | |
| RPS17S Forward | TCCGTGGTATCTCCATCAAGCT |
| RPS17S Reverse | CACTTCCGGCACGTAGTTGTC |
| RPS17S Probe | HEX/CAGGAGGAG/ZEN/GAACGTGAGCGCAG/3lABkFQ |
| Ae. aegypti kdr screening – qPCR | |
| 1016 Forward | CGTGCTAACCGACAAATTGTTTCC |
| 1016 Reverse | GACAAAAGCAAGGCTAAGAAAAGGT |
| 1016 Probe Val+ | VIC/CCGCACAGATACTTA/NFQ |
| 1016 Probe Ilekdr | FAM/CCCGCACAGGTACTTA/NFQ |
| 1534 Forward | CGAGACCAACATCTACATGTACCT |
| 1534 Reverse | GATGATGACACCGATGAACAGATTC |
| 1534 Probe Phe+ | FAM/ACGACCCGAAGATGA/NFQ |
| 1534 probe Cyskdr | VIC/AACGACCCGCAGATGA/NFQ |
| Wolbachia – qPCR | |
| WSPTM2 Forward | CATTGGTGTTGGTGTTGGTG |
| WSPTM2 Reverse | ACACCAGCTTTTACTTGACCAG |
| WSPTM2 Probe | FAM/TCCTTTGGA/ZEN/ACCCGCTGTGAATGA/3lAbRQSp |
| Wolbachia – LAMP | |
| FIP | TGTATGCGCCTGCATCAGCTTCGGTTCTTATGGTGCTAA |
| BIP | GCAGAAGCTGGAGTAGCGTTGTGTCATGCCACTTAGATGG |
| F3 | TGATGTAACTCCAGAAGTCA |
| B3 | CTTATTGGACCAACAGGATCG |
| LpF | AGCCTGTCCGGTTGAATT |
| LpB | CAGTCTTGTTATCCCAGTGAGT |

**Supplementary Figures**:

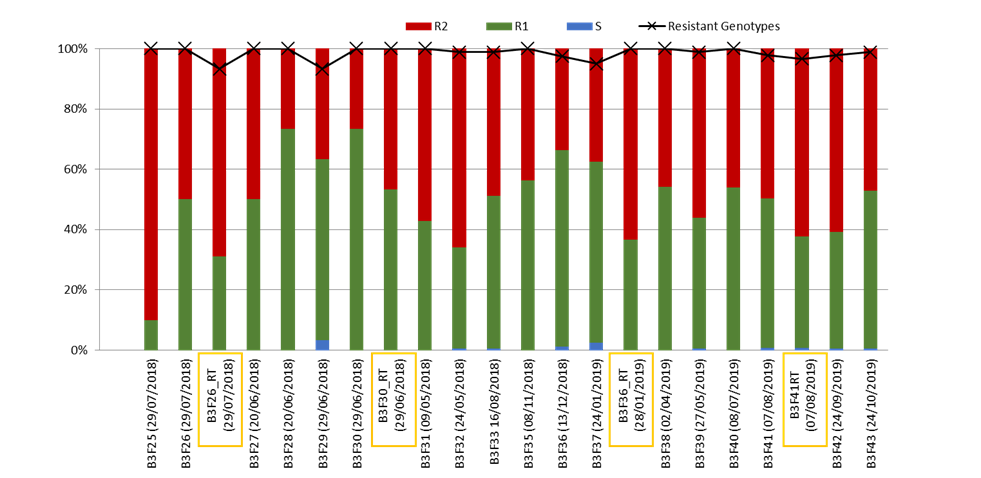

**Figure S1: KDR genotype analysis of consecutive wMelRio brood stock generations from F25 to F43 showing 4 outcrossing events.** Outcrossing events were performed on brood females in generation F26, F30, F36 and F41. ‘_RT’ represents the offspring of those outcrossing events and are marked with an orange box. Field collected samples in Rio and Niteroi always show highly resistant genotypes with a roughly 50:50 frequency distribution of R1 and R2 mutations. The wMelRio brood stock line tends to increase its R1 frequency with standard inbred rearing and some small % of susceptible genotypes start to appear around the 3rd Generation. The resulting outcross event normally restores the near 50:50 R1:R2 frequency distribution and reduces susceptible genotype frequencies as well. Methods for kdr genotyping, primers and probes are presented in supplementary methods.

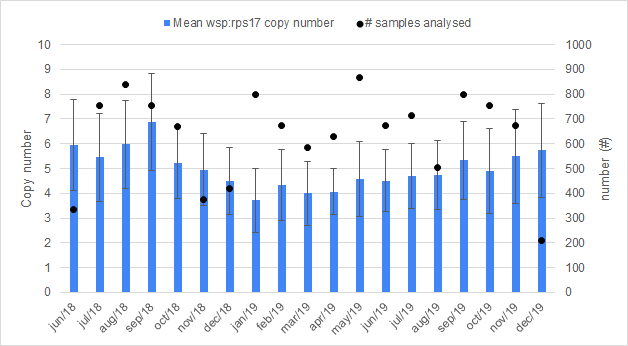

**Figure S2: Analysis of *w*Mel quantification in the release generation.** Quantification of *w*Mel was performed weekly on up to 4 day old mosquitoes from the release generation, emerged within the release device, prior to releases. *wsp:rps*17 copy numbers were fairly constant between 4 to 6, from June 2018 to December 2019. Error bars represent standard deviation of the mean. Total numbers of mosquitoes tested are represented by black dots.

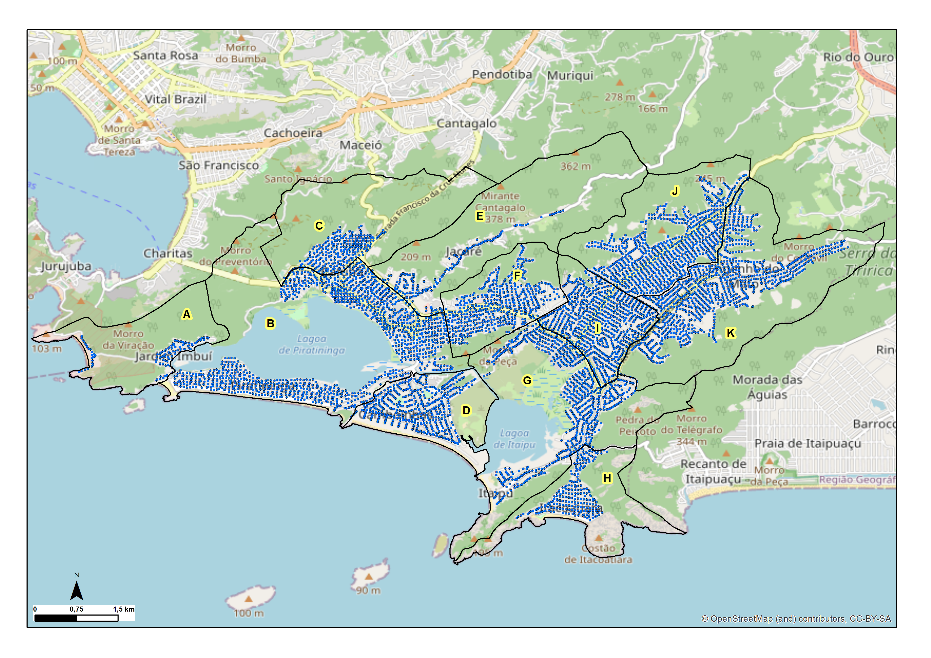

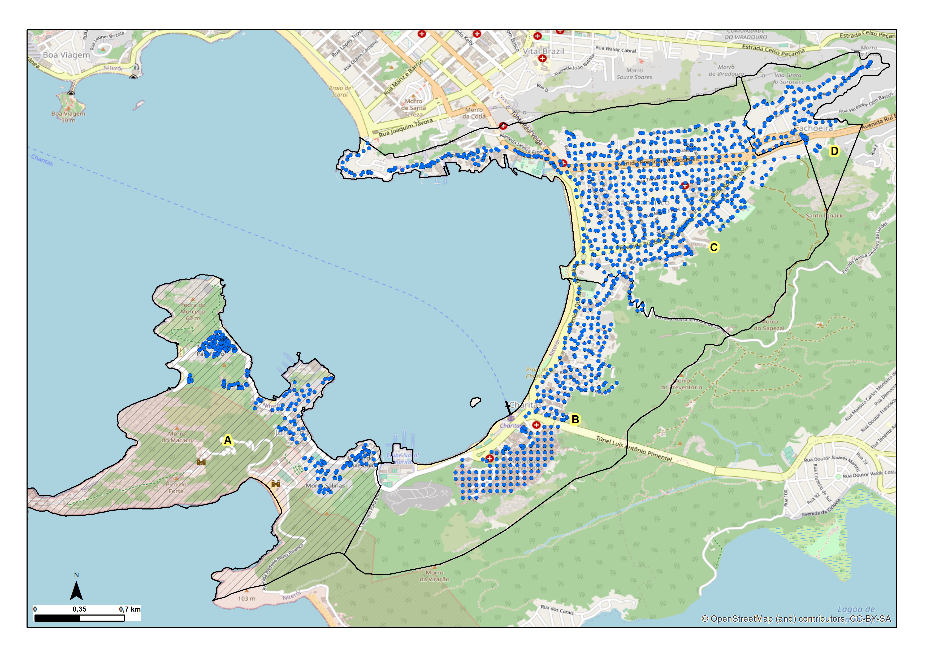

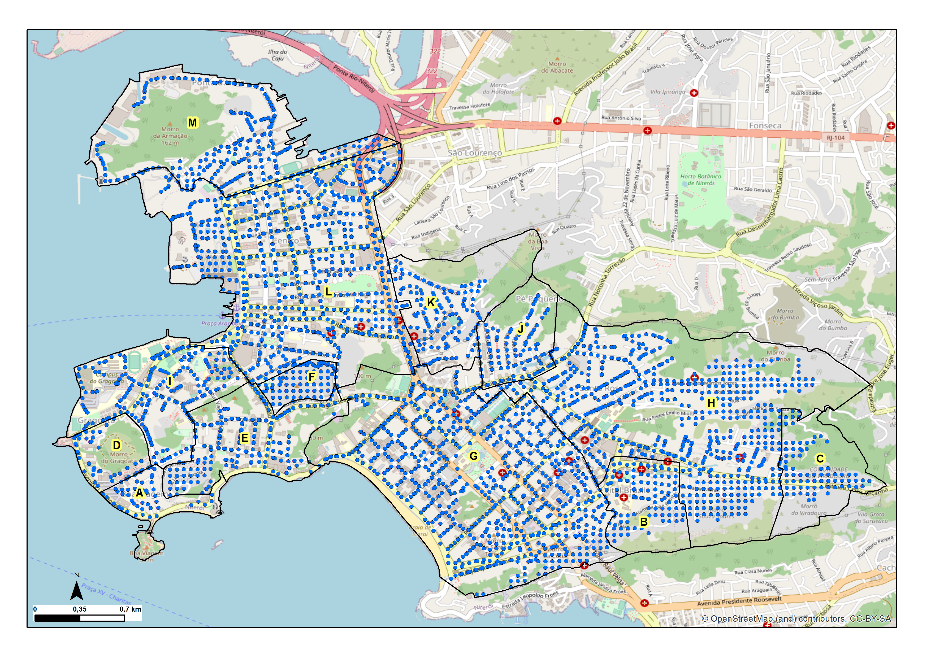

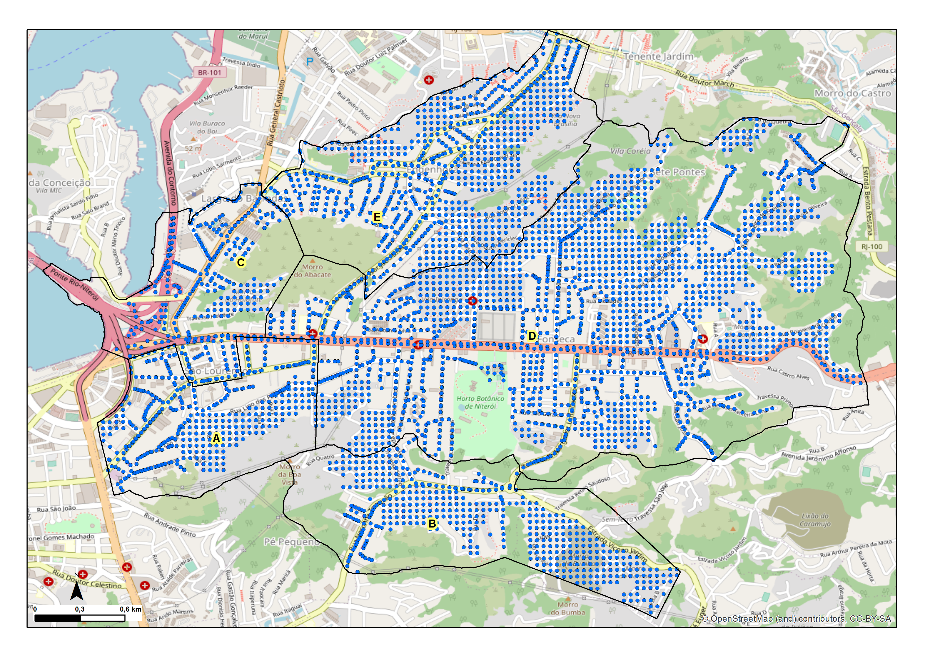

**Figure S3: Spatial distribution of mosquito release locations** in Niterói release zone 1 (A), zone 2 (B), zone 3 (C) and zone 4 (D)**.** Approximate locations of adult mosquito releases are shown by blue markers. The Jurujuba pilot release area in zone 1 is indicated with hatched shading. Maps were generated using ArcGIS 10.7 (Esri, Redlands, CA, USA) and ^©^OpenStreetMap source data.

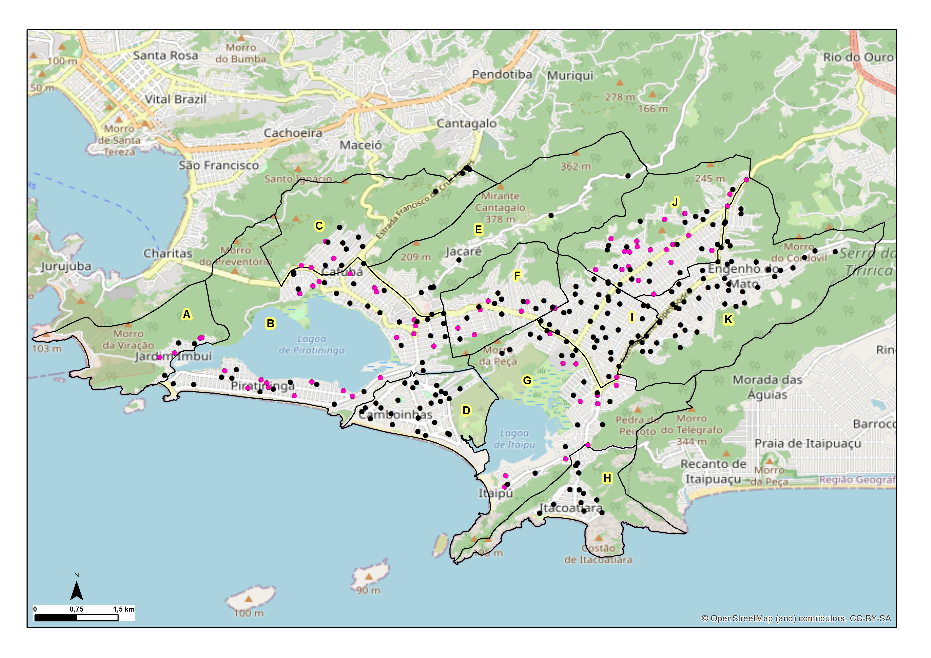

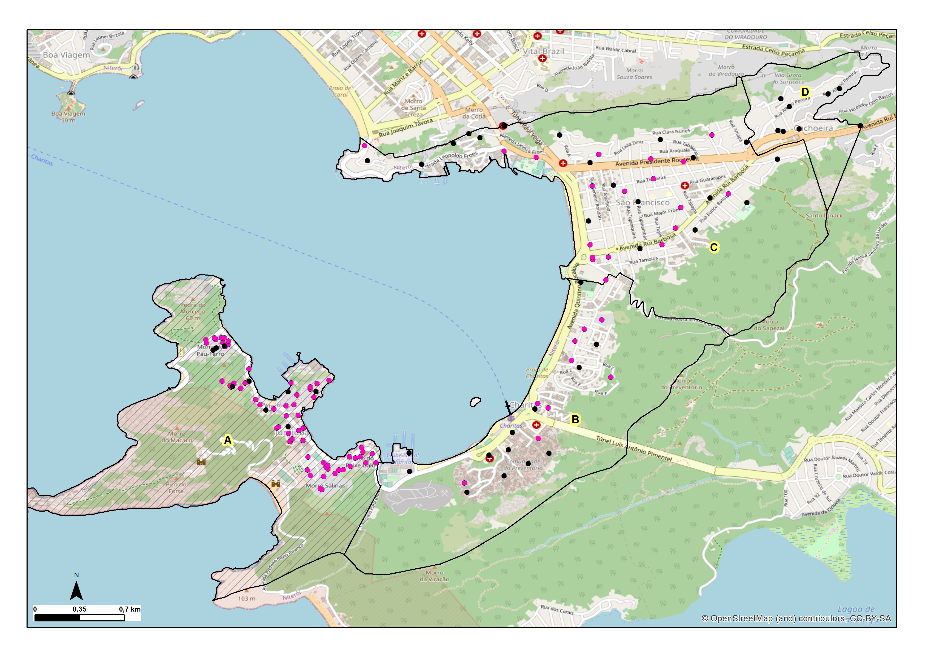

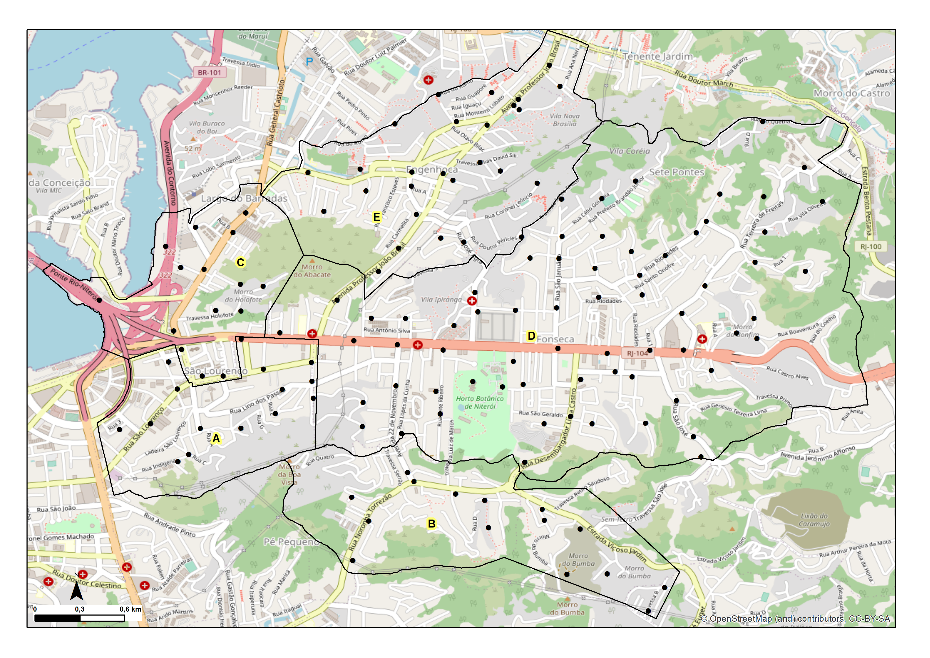

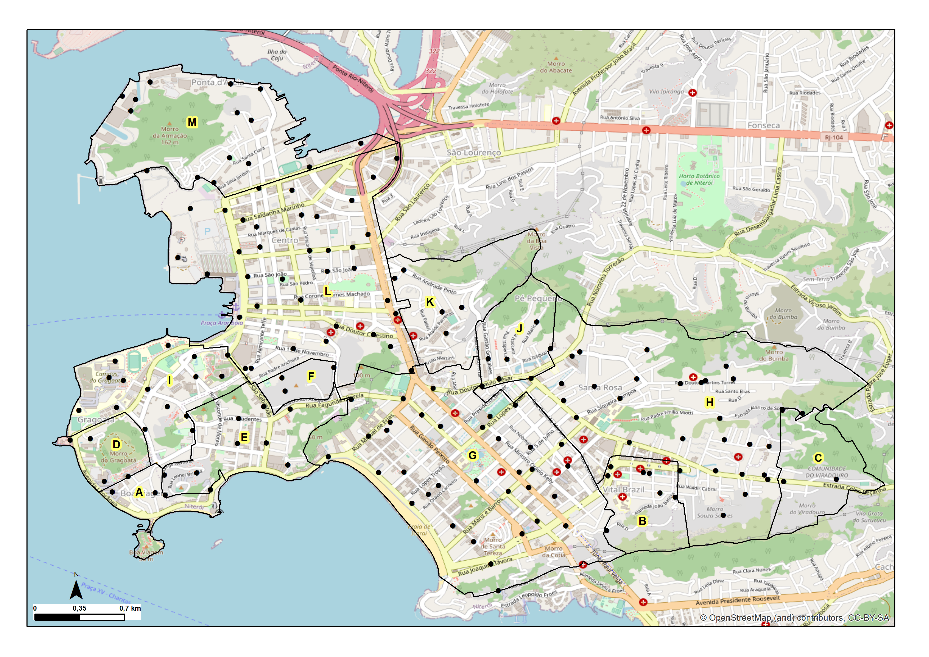

**Figure S4: Spatial distribution of mosquito monitoring locations** in Niterói release zone 1 (A), zone 2 (B), zone 3 (C) and zone 4 (D)**.** Approximate locations of BG adult mosquito traps are shown for each zone. Black markers indicate BG traps that were retained throughout the monitoring period. Pink markers indicate BG traps that were removed in three of four neighbourhoods in zone 1 and six of 11 neighbourhoods in zone 2 once releases were completed and *w*Mel prevalence was >60% in 3 consecutive monitoring events measured at least 4 weeks after the conclusion of releases, in order to reduce monitoring costs. The Jurujuba pilot release area in zone 1 is indicated with hatched shading. Maps were generated using ArcGIS 10.7 (Esri, Redlands, CA, USA) and ^©^OpenStreetMap source data

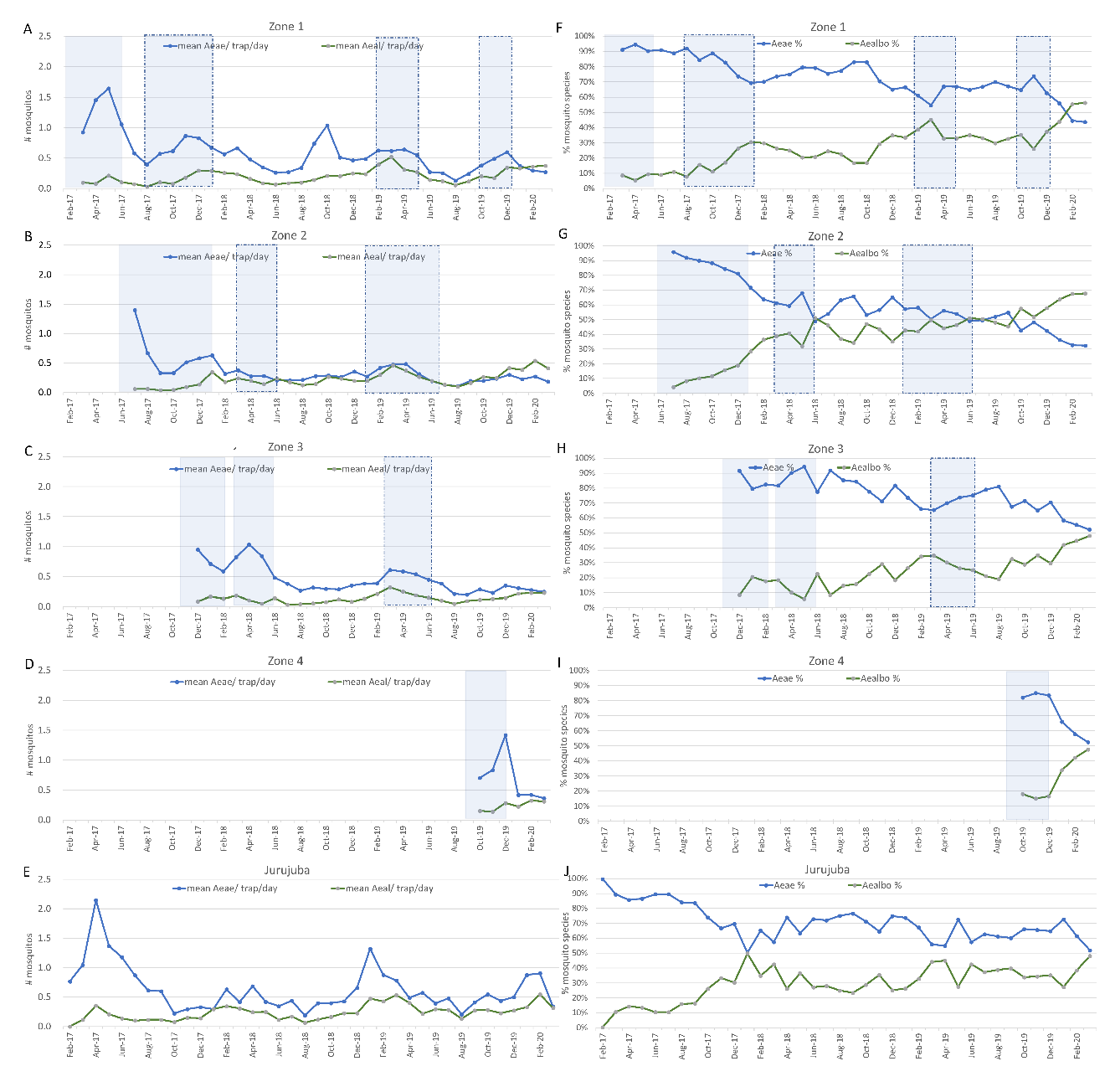

**Figure S5: Analysis of the abundance of *Aedes albopictus* and *Aedes aegypti* in BG trap collections in Niterói** **release zones.** Panels A–E show the mean number of mosquitoes caught per trap per day for each species, each month, in release zones 1–4 and in the Jurujuba pilot release area. Panels F–J show the relative frequency of each species, each month, in release zones 1–4 and in the Jurujuba pilot release area. Shaded areas represent release periods. Dotted shaded areas indicate that only part of the zone was receiving releases.

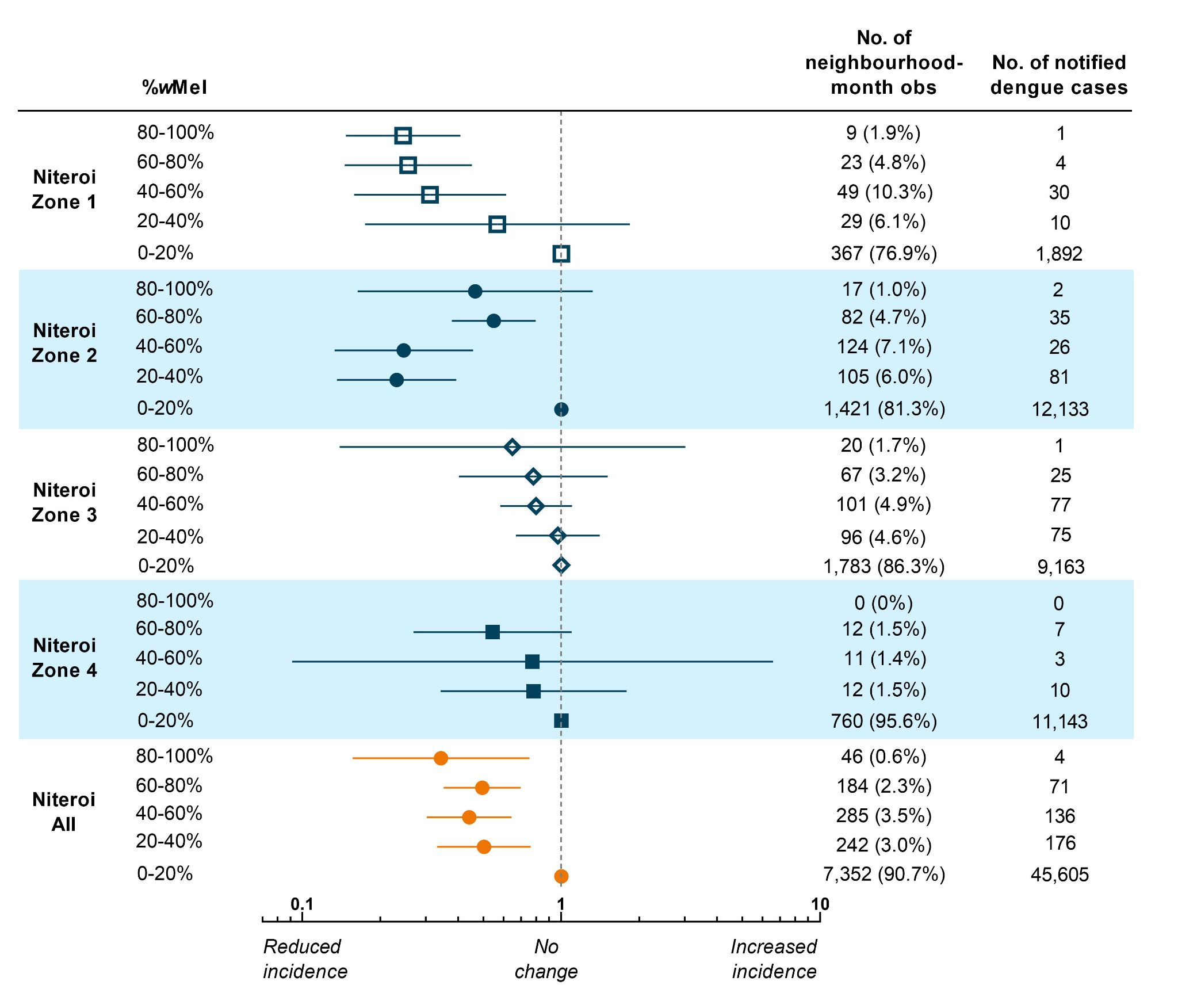

**Figure S6: Estimated reduction in dengue incidence with increasing *w*Mel prevalence in *Aedes aegypti* populations in Niterói neighbourhoods.** This analysis uses a three-month moving average of *w*Mel% and excludes the Zone 1 pilot release area of Jurujuba. Point estimates (markers) and 95% confidence intervals (horizontal bars) are from controlled interrupted time series analysis of monthly dengue case notifications to the Brazilian national disease surveillance system (Jan 2007 – March 2020), by neighbourhood, in each release zone and in the aggregate release area. *w*Mel prevalence was calculated as the percentage of trapped *Ae. aegypti* positive for wMel, in each neighbourhood each month, grouped by quintile. The lowest quintile (*w*Mel 0-20%) served as the reference category for calculation of the incidence rate ratio (IRR) and included the monthly observations within that quintile from the respective release zone, as well as all observations from the untreated control zone (n=3,021 neighbourhood-months observed and n=11,278 notified dengue cases).

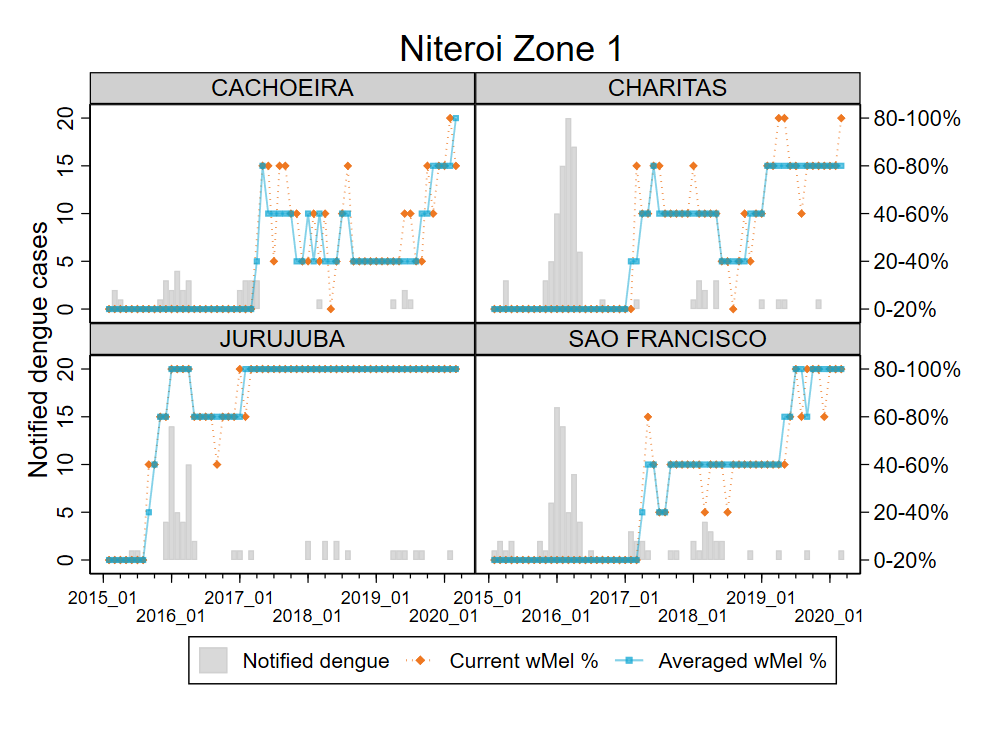

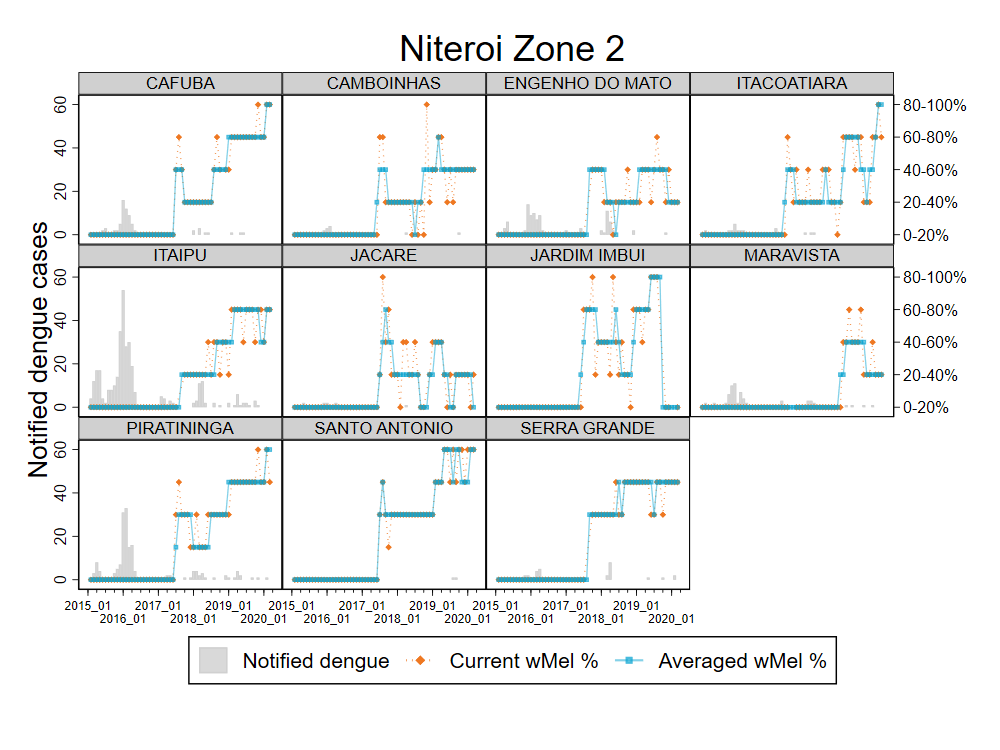

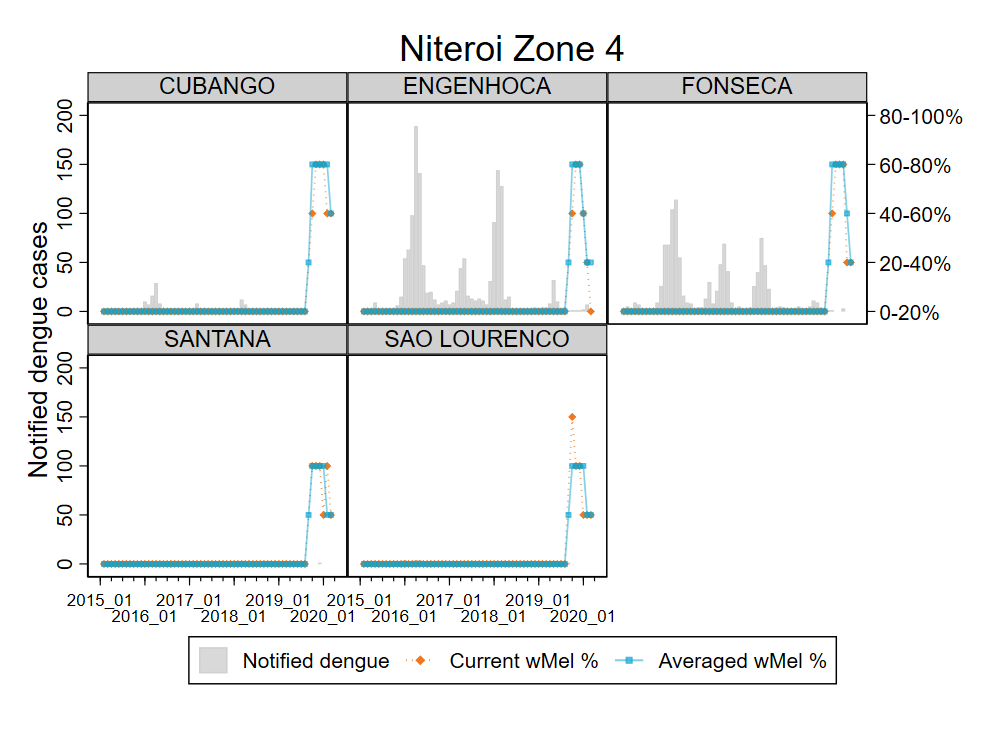

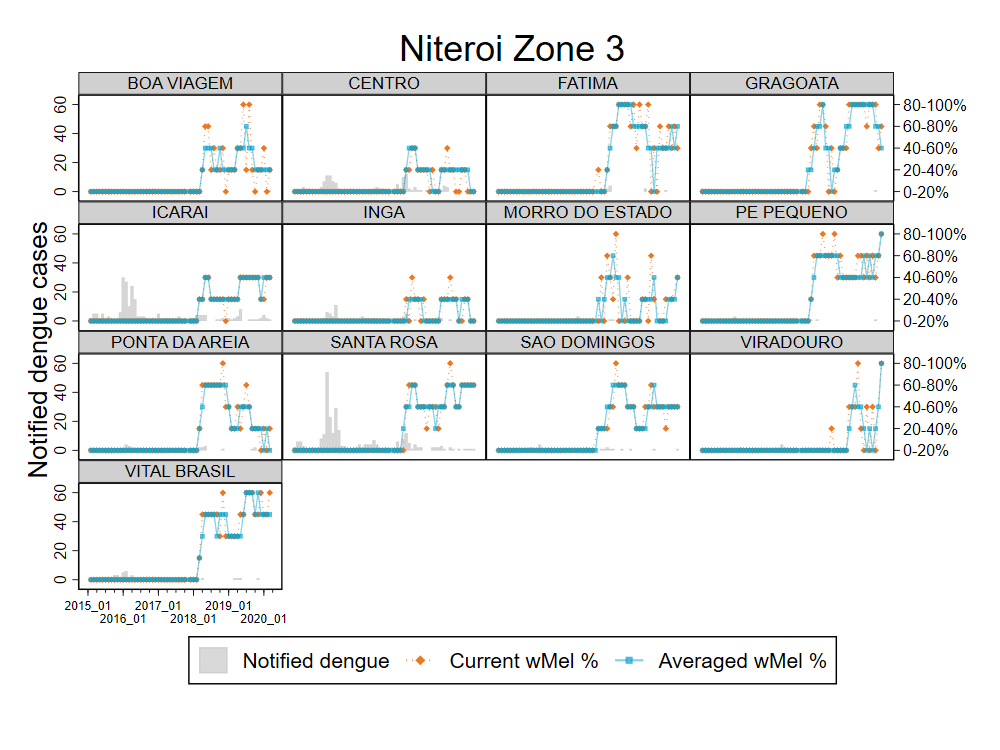

**Figure S7: Notified dengue cases and *w*Mel% quintile monthly time series by neighbourhood, in Niterói release zones 1 – 4.** *w*Mel% quintile was based on the *w*Mel prevalence in a single month (current *w*Mel%) or a three-month moving average (Averaged *w*Mel%).

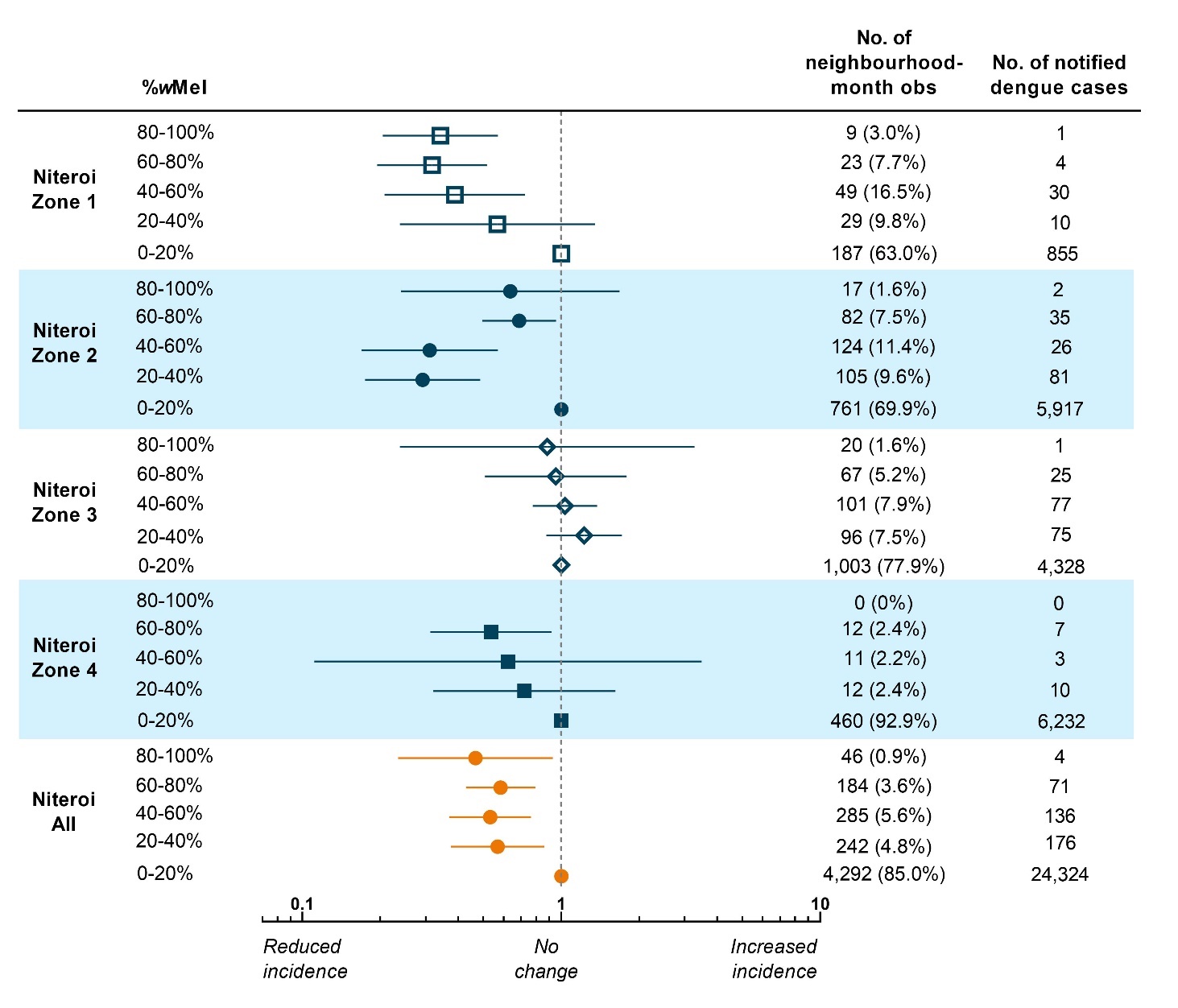

**Figure S8: Estimated reduction in dengue incidence with increasing *w*Mel prevalence in *Aedes aegypti* populations in Niterói neighbourhoods – sensitivity analysis.** This sensitivity analysis	excludes	all observations prior to 2012, five years prior to the start of releases. Point estimates (markers) and 95% confidence intervals (horizontal bars) are from controlled interrupted time series analysis of monthly dengue case notifications to the Brazilian national disease surveillance system (Jan 2012 –	March 2020), by neighbourhood, in each release zone and in the aggregate release area. *w*Mel prevalence was calculated as the percentage of trapped *Ae. aegypti* positive for *w*Mel, in each neighbourhood in a moving three-month window, grouped by quintile. The lowest quintile (*w*Mel 0-20%) served as the reference category for calculation of the incidence rate ratio (IRR) and included the observations within that quintile from the respective release zone, as well as all observations from the untreated control zone (n=1,881 neighbourhood-months observed and n=6,996 notified dengue cases).

**Supplementary Tables**:

**Table S1. Dengue, chikungunya and Zika incidence rate ratios in *Wolbachia*-release zones compared to the control zone.** IRRs are from negative binomial regression models of monthly case counts (Jan 2007 – June 2020 for dengue; Jan 2015 – June 2020 for chikungunya and Zika), with an offset for population size and 6-monthly flexible cubic splines to account for seasonal effects. The mixed effects model for the aggregate Niteroi release area included a random effect for release zone.

|  | Incidence rate ratio (95% confidence interval) | | |
| --- | --- | --- | --- |
|  | Dengue | Chikungunya | Zika |
| Zone 1 | 0.30 (0.19, 0.47) | 0.30 (0.15, 0.59) | 0.44 (0.18, 1.09) |
| Zone 2 | 0.24 (0.15, 0.38) | 0.93 (0.50, 1.73) | 0.66 (0.26, 1.63) |
| Zone 3 | 0.54 (0.37, 0.79) | 0.30 (0.17, 0.53) | 1.04 (0.50, 2.16) |
| Zone 4 | 0.31 (0.18, 0.54) | 0.29 (0.13, 0.67) | 0.60 (0.17, 2.05) |
| Niteroi | 0.31 (0.21, 0.46) | 0.44 (0.23, 0.84) | 0.63 (0.40, 0.99) |

**Table S2. Dengue incidence rate ratios with increasing *w*Mel prevalence in *Aedes aegypti* populations in Niteroi neighbourhoods.** IRRs are from mixed effects negative binomial regression models of monthly dengue case counts (Jan 2007 – March 2020) by neighbourhood, with an offset for population size, 6-monthly flexible cubic splines to account for seasonal effects, and a random effect for neighbourhood.

|  | Incidence rate ratio (95% confidence interval) | | | | |
| --- | --- | --- | --- | --- | --- |
| *w*Mel% quintile | Zone 1 | Zone 2 | Zone 3 | Zone 4 | Niteroi |
| 0-20% | Ref | Ref | Ref | Ref | Ref |
| 20-40% | 0.56  (0.17, 1.83) | 0.23  (0.13, 0.39) | 0.96  (0.66, 1.40) | 0.78  (0.34, 1.78) | 0.50  (0.33, 0.76) |
| 40-60% | 0.31  (0.15, 0.61) | 0.24  (0.13, 0.45) | 0.79  (0.58, 1.09) | 0.77  (0.09, 6.57) | 0.44  (0.30, 0.64) |
| 60-80% | 0.25  (0.14, 0.45) | 0.54  (0.37, 0.79) | 0.77  (0.40, 1.51) | 0.54  (0.26, 1.09) | 0.49  (0.35, 0.69) |
| 80-100% | 0.24  (0.14, 0.40) | 0.46  (0.16, 1.32) | 0.64  (0.13, 3.01) | - | 0.34  (0.15, 0.75) |

**Table S3. Dengue incidence rate ratios with increasing *w*Mel prevalence in *Aedes aegypti* populations in Niteroi neighbourhoods – sensitivity analysis** excluding pre-intervention observations prior to 2012 to achieve greater balance in the length of pre-intervention and post-intervention observation periods. IRRs are from mixed effects negative binomial regression models of monthly dengue case counts (Jan 2012 – March 2020) by neighbourhood, with an offset for population size, 6-monthly flexible cubic splines to account for seasonal effects, and a random effect for neighbourhood.

|  | Incidence rate ratio (95% confidence interval) | | | | |
| --- | --- | --- | --- | --- | --- |
| *w*Mel% quintile | Zone 1 | Zone 2 | Zone 3 | Zone 4 | Niteroi |
| 0-20% | Ref | Ref | Ref | Ref | Ref |
| 20-40% | 0.56  (0.23, 1.34) | 0.29  (0.17, 0.48) | 1.22  (0.87, 1.71) | 0.71  (0.32, 1.61) | 0.56  (0.37, 0.85) |
| 40-60% | 0.38  (0.20, 0.72) | 0.31  (0.16, 0.56) | 1.03  (0.77, 1.37) | 0.62  (0.11, 3.48) | 0.53  (0.36, 0.76) |
| 60-80% | 0.31  (0.19, 0.51) | 0.68  (0.49, 0.95) | 0.95  (0.50, 1.78) | 0.53  (0.31, 0.91) | 0.58  (0.42, 0.79) |
| 80-100% | 0.34  (0.20, 0.56) | 0.63  (0.24, 1.67) | 0.88  (0.23, 3.26) | - | 0.46  (0.23, 0.92) |

**Supplementary References:**

1.	Hayd RLN, Carrara L, de Melo Lima J, de Almeida NCV, Lima JBP, Martins AJ. Evaluation of resistance to pyrethroid and organophosphate adulticides and kdr genotyping in *Aedes aegypti* populations from Roraima, the northernmost Brazilian State. Parasit Vectors 2020;13:264.

2.	Macoris ML, Martins AJ, Andrighetti MTM, Lima JBP, Valle D. Pyrethroid resistance persists after ten years without usage against *Aedes aegypti* in governmental campaigns: Lessons from Sao Paulo State, Brazil. PLoS Negl Trop Dis 2018;12:e0006390.
